# Chronic early-life obesity linked to childhood impulsivity predicts long-term psychosis trajectory through dose-dependent cerebellar dysmaturation in 22q11.2 Deletion Syndrome

**DOI:** 10.1101/2025.02.13.25322209

**Authors:** Corrado Sandini, Natacha Reich, Farnaz Delavari, Lara Pajic, Andrea Escelsior, Silas Forrer, Andrea Imparato, Nada Kojovic, Caren Latreche, Valeria Parlatini, Samuele Cortese, Maude Schneider, Stephan Eliez

**Author notes:** **Corresponding author:** Corrado Sandini, Developmental Imaging and Psychopathology Laboratory, Department of Psychiatry, University of Geneva School of Medicine, Campus Biotech, Chemin des Mines 9, 1202 Geneva, Switzerland.

## Abstract

**Background:** Recent epidemiological evidence links early-life obesity and metabolic dysregulation to adult psychosis vulnerability, though a causal relationship remains unclear. Establishing causality in highly heritable psychotic disorders requires: 1) demonstrating that early-life metabolic factors mediate between genetic vulnerability and psychosis trajectory, 2) dissecting mechanisms leading to early-life obesity in genetically vulnerable individuals, and 3) clarifying downstream neurodevelopmental pathways linking early-life obesity to psychosis symptoms.

**Methods:** To address these key issues, we investigated bidirectional pathways linking behavioral, BMI, and neurodevelopment trajectories in a unique longitudinal cohort of 184 individuals at high genetic risk for psychosis, due to 22q11.2 Deletion Syndrome (22q11DS), and 182 neurotypical controls, followed-up since childhood. We combined repeated BMI measurements with clinical/neurocognitive phenotyping and neuroimaging. We investigated the relationship between BMI trajectories with risk of psychosis and tested whether altered cortical or cerebellar development could underlie this association.

**Results:** Childhood behavioral impulsivity predicted early and progressive deviations in BMI trajectories. In turn, pubertal BMI-increases were associated with concomitant worsening of depressive symptoms, while chronic BMI-increases emerging during childhood predicted the subsequent emergence of psychosis during late-adolescence/early-adulthood. The duration of increased BMI-status was associated with emergence of motor and cognitive disorganization, a key schizophrenia symptom domain, which was linked to progressive gray matter volume reductions in posterior-inferior cerebellum.

**Conclusions:** These findings suggest that metabolic dysregulation associated with obesity may link childhood behavioral impulsivity to psychosis vulnerability in 22q11DS, by influencing cerebellar maturation. These findings might support preventive interventions targeting early-life metabolic trajectories in individuals at risk of psychosis.

## Introduction

Obesity is significantly related to multiple forms of psychopathology, including neurodevelopmental, mood, and psychotic disorders (1). The causal directionality and mechanisms accounting for these associations remain largely unclear. Indeed, metabolic dysregulation associated with obesity could contribute to the emergence of psychopathology, or be the result of altered caloric intake and expenditure in the context of psychiatric disorders (1, 2), or their treatment (3). To clarify the direction of these associations, Perry et al. investigated metabolic trajectories in a large epidemiological sample of over 14000 individuals, followed-up longitudinally from birth until 24 years of age (4). Results suggested that a progressive BMI increase emerging during childhood was associated with increased fasting insulin and inflammatory markers persisting throughout adolescence, which ultimately contributed to a 6-fold increased risk of developing a psychotic disorder in early-adulthood (4). These findings support a causal link between childhood obesity/metabolic dysregulation and adult psychosis vulnerability, mediated by a prolonged pro-inflammatory state throughout adolescence. If confirmed this would carry significant clinical implications, given that metabolic trajectories can be effectively targeted through a variety of therapeutic and preventive public-health strategies (5). Currently however, at least 3 key steps in the causality chain remain insufficiently clear (2) (4).

Firstly, although psychosis is highly heritable (6), existing studies have still not demonstrated that early-life obesity mediates the link between genetic vulnerability and psychosis clinical trajectories (2). The 22q11.2 Deletion Syndrome (22q11DS) represents a unique genetic model to dissect the long-term developmental mechanisms contributing to the emergence of psychosis (7). The genetic alterations associated with 22q11DS represent the single strongest genetic risk factor for psychosis, with an adult prevalence of psychotic disorders of 30-40% (8). Moreover 22q11DS also contributes vulnerability to atypical early-life BMI trajectories ultimately leading to a high prevalence of adult obesity (9). Currently, the association between behavioral and metabolic phenotypes of 22q11DS remains largely unclear. Here we investigated for the first time whether atypical BMI trajectories would contribute to psychosis vulnerability in a cohort of 184 genetically vulnerable individuals with 22q11DS and 182 healthy controls, followed-up longitudinally from childhood to adulthood (for a total of 756 longitudinal assessments).

Secondly, it is still unclear which mechanism might contribute to early-life obesity in individuals at genetic risk for psychosis (2). Interestingly, although the pathophysiology of early-life obesity remains partially understood(10), both clinical and genetic evidence (11, 12) increasingly implicate the role behavioral factors (13). In particular, the hyperactivity/impulsivity dimensions of Attention-Deficit/Hyperactivity Disorder (ADHD), is a highly replicated risk factor for early-life obesity(14), which is directly corrected by ADHD treatment (14, 15), suggesting a disease specific causal mechanism (16). It was proposed that this association might be partially explained by the impact behavioral impulsivity on obesity-predisposing eating patterns(17). Of note, 22q11DS is associated with 30% of ADHD during childhood (8). Here we investigated whether behavioral impulsivity associated with ADHD would predispose to deviant BMI trajectories in 22q11DS. We hypothesized that this behavioral-metabolic pathway could contribute to account for the 3-way genetic and clinical association between ADHD, early-life obesity and psychosis, observed both in 22q11DS and in the general population (4, 18).

Thirdly, it remains unclear whether early-life obesity directly influences downstream neurodevelopmental trajectories implicated in the emergence of psychosis symptoms, through a dose-dependent effect (2). Of note, bot obesity and psychosis are associated with significant reductions of Gray-Matter-Volume (GMV) which, while not identical in their anatomical distribution, overlap in affecting Cerebellar-GMV (19, 20). Immune dysregulation has been suggested to play a central role in reductions in gray-matter volume, observed in both obesity (21) and psychosis(22). Indeed, metabolic syndrome associated with obesity has been identified as one of the strongest risk factors for the development of chronic pro-inflammatory states (23), that have been strongly implicated in the pathophysiology of psychosis (2, 22), possibly due to their negative effects on synaptic pruning during adolescence (24). Intriguing evidence has recently suggested that obesity and immune dysregulation might account for gray-matter volume reductions in first-episode psychosis (25), but the longitudinal studies required to dissect such 3-way interactions have not yet been conducted. Here we modelled longitudinal development of cerebellar and cortical gray-matter volume from childhood to adulthood, as function of both early-life BMI and psychiatric trajectories in individuals with 22q11DS. We hypothesized that prolonged early-childhood obesity would exert a duration-dependent effect on gray-matter volume reductions, in particularly impacting the cerebellum, that would be specifically implicated in the emergence of disorganization symptoms of psychosis. Overall, we hypothesized the long-term multimodal phenotyping of individuals with 22q11DS would provide a unique view of developmental mechanisms linking ADHD-related behavioral impulsivity, to early-life obesity and subsequent psychosis vulnerability.

## Methods

### Participants

In the present study, 372 participants, encompassing 184 participants with 22q11DS (M/F=93:91) and 188 Healthy Controls (HC) (M/F=92:96), were recruited as part of the Geneva-22q11DS Longitudinal Study (26) and followed-up for a total of 765 longitudinal assessments (433 in 22q11DS and 333 in HC). Table 1 reports participants’ characteristics. Most (146/198) HC were recruited among unaffected siblings of 22q11DS participants.

**Table 1:**
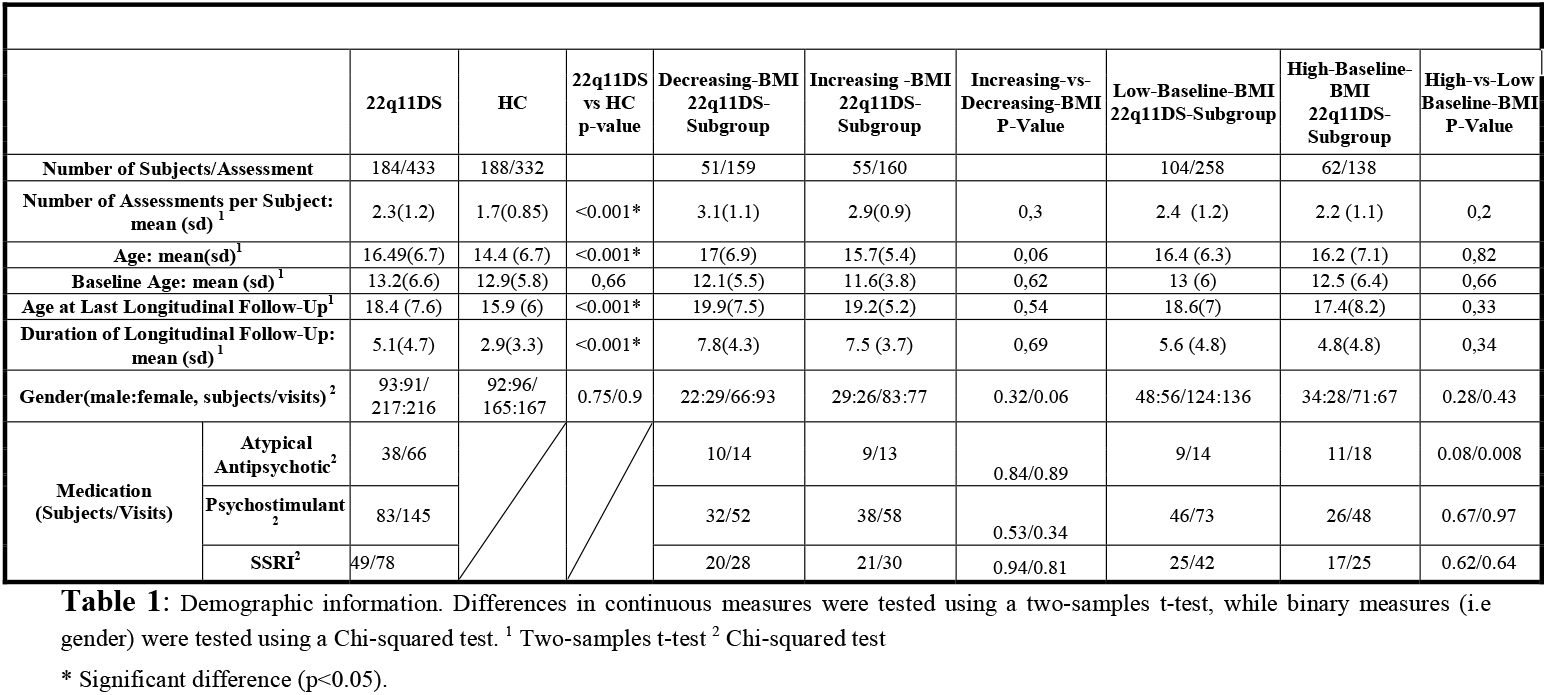
Part 1: Demographics Features.

### Clinical and neurocognitive measures

Psychiatric diagnoses were assessed according to the DSM-5 (APA, 2013) with a combination of age-adapted semi-structured interviews detailed in the supplementary material. To further characterize clinical trajectories, we combined the Structured Interview for Prodromal Syndromes (SIPS) (27), the Brief Psychiatric Rating Scale (BPRS) (28), and the Child/Adult Behavior Checklist (CBCL/ABCL) (29). Neurocognitive impulsivity was measured with the Conners’ Continuous Performance Test 2^nd^ edition (CPT2), considering age-normed T-Scores of Hit-Reaction-Time and the T-Score-ratio of Commission/Omission errors (30). Intelligent quotient (IQ) was measured using the Wechsler Intelligence Scale for Children (WISC-IV or V)(31) and Adults (WAIS-III or IV) (32)).

### Statistical Analyses

Statistical analyses were conducted in MATLAB (v-2021a). We first compared BMI trajectories between 22q11DS and HCs using mixed-model linear regression (MMLR), implemented via a previously published toolbox (https://github.com/danizoeller/myMixedModelsTrajectories. To control for potential treatment effects, we repeated this analysis including only antipsychotic-naïve participants. To explore clinical correlates by BMI, we categorized individuals with 22q11DS based on their BMI trajectories using a published pipeline (26). We defined average BMI trajectories for males and females, then classified them based on whether their baseline BMI was above or below the age-equivalent gender-specific trajectory. This procedure was repeated for BMI rate-of-change during longitudinal follow-up, considering only antipsychotic-naïve assessments. This yielded High-vs-Low Baseline-BMI and Increasing-vs-Decreasing BMI-Trajectory subgroups (see Table 1). We used MMLR to compare subgroups regarding clinical and neurocognitive trajectory development, applying False Discovery Rate correction for multiple comparisons across psychometric instruments. For discrete lifetime outcomes, such as psychiatric diagnoses or psychotropic medication use, we computed Kaplan-Meier survival curves. Finally, we assessed whether clinical outcomes were better explained by the duration of High-BMI status rather than other age-related factors, modeling clinical trajectories as a function of Time from Baseline Assessment (TBA) across Baseline-BMI subgroups.

### Structural MRI Image Acquisition, Processing and Analysis

T1-weighted images were acquired using Siemens Trio or Prisma 3T scanners (see Supplementary Material for details). Cerebellar segmentation followed the CERES method, implemented via the Volbrain MRI volumetry platform(33). Gray Matter Volume (GMV) measures were extracted for the entire cerebellum and 12 lobules, averaged across hemispheres. Cortical segmentation was performed using Freesurfer (Version 7.4.1), with GMV extracted from 34 cortical regions (34) and averaged across hemispheres. High-quality cortical and cerebellar segmentation was available for 139 individuals with 22q11DS (254 assessments).

Cortical and cerebellar GMV trajectories were compared across BMI subgroups using MMLR, controlling for scan type, gender, psychotropic medication use, and intracranial volume. Additionally, we examined whether cerebellar trajectories linked to baseline BMI were better predicted by the duration of High-BMI status than by age, suggesting a possible dose-effect relationship.

Then, we explored 3-way associations between BMI status, neurodevelopmental trajectories, and clinical outcomes. Firstly, we investigated whether atypical GMV trajectories linked to BMI-Status were also associated to the emergence of disorganized communication, odd behavior, bizarre thinking, or motor disturbances of moderate/severe intensity. These symptoms were selected due to their strong association with prolonged high-BMI status. Secondly, we performed Partial Least Squares Correlation (PLS multivariate) analysis (https://github.com/MIPLabCH/myPLS), blind to BMI-status, to dissect associations between multivariate clinical patterns and GMV trajectories. See supplementary material for details. Significant associations between clinical and neurodevelopmental trajectories were summarized in composite clinical-neurodevelopmental scores computed at each longitudinal assessment. We compared the trajectories of such scores across BMI subgroups, to assess whether BMI status contributed to the multivariate clinical-neurodevelopmental associations detected by PLSC.

## Results

### Behavioral impulsivity predicts divergent BMI trajectories in 22q11DS

In both controls and individuals with 22q11DS, BMI followed a cubic trajectory, increasing steeply during adolescence and stabilizing in early adulthood (Figure-1-Panel-1A). However, individuals with 22q11DS exhibited a significantly steeper BMI increase from late childhood/early adolescence (P-Age-Interaction<0.001), leading to a higher BMI than controls by late adolescence/early adulthood (P-Group-Effect<0.001). BMI increases preceded antipsychotic medication use and remained significant even in antipsychotic-naïve assessments (Figure-1-Panel-1B).

**Figure 1:**
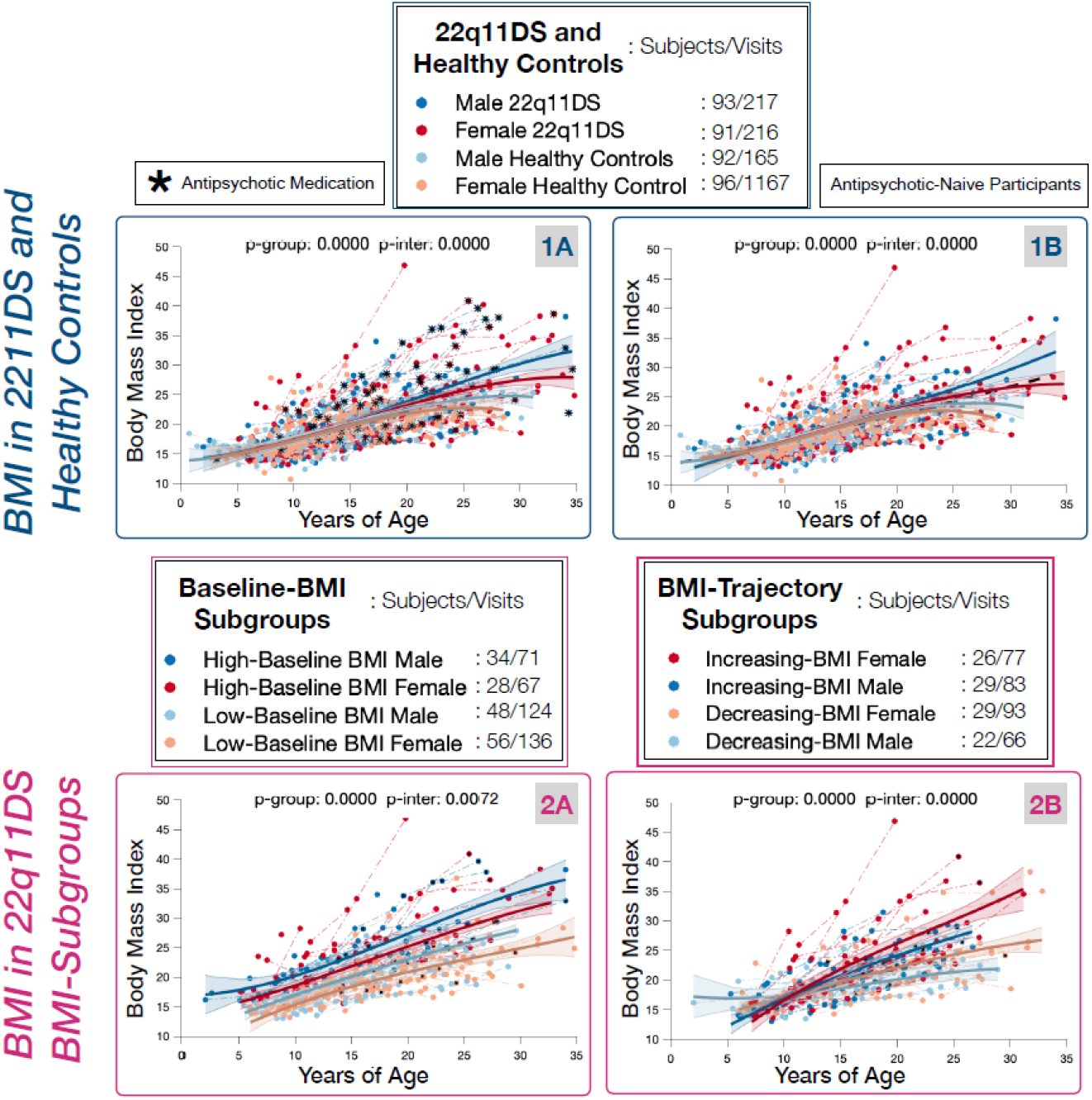
**Panel 1A:** Developmental trajectories of BMI scores compared across Healthy Controls and 22q11DS individuals modelling for the effects of gender yielding 4 subgroups: Male 22q11DS in dark blue, Female 22q11DS in red, Male Healthy Controls in Light Blue, Female Healthy Controls in Orange. Assessments during which 22q11DS individuals are receiving antipsychotic treatment are highlighted in black. **Panel 1B:** Developmental trajectories of BMI scores compared across Healthy Controls and 22q11DS individuals still naïve to antipsychotic medication. **Panel 2A:** Subgroup analysis within 22q11DS sample. 22q11DS are divided in subgroups according to BMI scores at baseline assessments and considering males and females separately. This analysis yields 4 subgroups, High-Baseline BMI-Males in dark blue, High-Baseline-BMI-Females in red, Low-Baseline-BMI-Males in light blue, and Low-Baseline-BMI-Females in orange. **Panel 2B:** Subgroup analysis within 22q11DS sample. 22q11DS are divided in subgroups according longitudinal BMI trajectories, considering males and females separately. This analysis yields 4 subgroups, Increasing-BMI-Males in dark blue, Increasing-BMI-Females in red, Decreasing-BMI-Males in light blue, and Decreasing-BMI-Females in orange.

BMI-Trajectory-Subgroups within the 22q11DS sample had similar baseline BMI but diverged in longitudinal trajectories (P-Age-Interaction<0.001), with the Female-Increasing-BMI-Subgroup showing the highest adult BMI (P-Group-Effect<0.0019) (Figure-1-Panel-2B). Baseline-BMI-Subgroups differed in BMI scores from childhood (P-Group-Effect<0.001), with the Male-High-Baseline-BMI-Subgroup showing the most pronounced divergence over time (P-Age-Interaction<0.001) (Figure-1-Panel-2A).

ADHD with combined impulsivity (≥3 DSM-5 Impulsivity/Hyperactivity symptoms) was significantly more frequent in both the High-Baseline-BMI (P-Group-Effect=0.0078) and Increasing-BMI-Subgroups (P-Group-Effect=0.0038), particularly in Male-Increasing-BMI (55%) and Female-High-Baseline-BMI (47%) subgroups, while lowest in Female-Low-Baseline-BMI (10%) and Female-Decreasing-BMI (8.9%) subgroups (Figure-2-Panels-1A/1B). Purely inattentive ADHD rates were similar across BMI subgroups (Supplementary-Figures-3/4). DSM-5 Impulsivity/Hyperactivity symptoms were significantly increased in High-Baseline-BMI (P-Group-Effect<0.0001, P-Age-Interaction<0.0001) and Increasing-BMI-Subgroups (P-Group-Effect=0.022, P-Age-Interaction=0.015), whereas DSM-5 Inattention symptoms did not differ (Figure-2-Panels-2A/2B, Supplementary Material).

**Figure 2:**
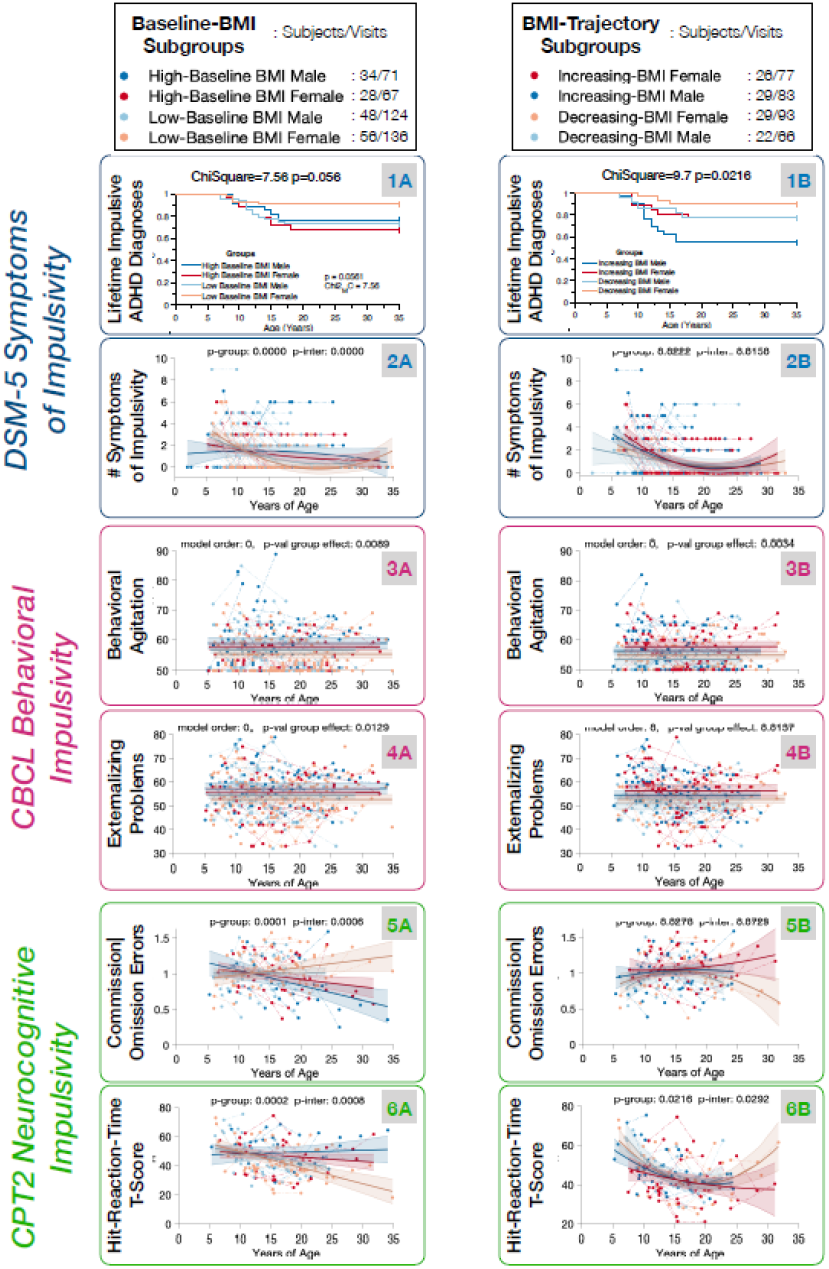
Evidence that atypical BMI trajectories are linked to childhood behavioral and neurocognitive impulsivity. **Panels 1A-6A:** Behavioral and neurocognitive impulsivity in High-vs-Low-Baseline-BMI subgroups. High-Baseline BMI-Males in dark blue, High-Baseline-BMI-Females in red, Low-Baseline-BMI-Males in light blue, and Low-Baseline-BMI-Females in orange. **Panels 1B-6B:** Behavioral and neurocognitive impulsivity in Increasing-vs-Decreasing-BMI-Trajectory subgroups. Increasing-BMI-Males in dark blue, Increasing-BMI-Females in red, Decreasing-BMI-Males in light blue, and Decreasing-BMI-Females in orange. **Panels 1A-1B:** Kaplan-Meyer survival curves for lifetime DSM-5 diagnosis of ADHD associated with 3 or more impulsivity/hyperactivity symptoms. **Panels 2A-2B:** Number of DSM-5 impulsivity/hyperactivity symptoms compared across BMI-Subgroups with Mixed-Model-Linear-Regression. **Panels 3A-3B:** Parentally reported Behavioral Agitation considering age-normed Child-Behavioral-Checklist T-Score compared across BMI-Subgroups with Mixed-Model-Linear-Regression. **Panels 4A-4B:** Parentally reported Externalizing Problems considering age-normed Child-Behavioral-Checklist T-Score compared across BMI-Subgroups with Mixed-Model-Linear-Regression. **Panels 5A-5B:** Ratio of CPT2 Commission/Omission Error T-Scores measuring neurocognitive impulsivity compared across BMI-Subgroups with Mixed-Model-Linear-Regression. **Panels 6A-6B:** CPT2 Hit-Reaction-Time T-Scores measuring neurocognitive impulsivity compared across BMI-Subgroups with Mixed-Model-Linear-Regression.

Parentally reported behavioral impulsivity was elevated in High-Baseline-BMI and Increasing-BMI-Subgroups, as shown by higher CBCL-Externalizing-Problems (P=0.0078) and CBCL-Behavioral-Agitation (P=0.0078) scores (Figure-2-Panels-2/3 and Supplementary-Figures-7/11). The High-Baseline-BMI subgroup also exhibited mild increases in CBCL-Internalizing-Problems, including anxiety, though anxiety disorder rates did not significantly differ across BMI subgroups (Supplementary-Figures-3/11).

Neurocognitive impulsivity in childhood was evident in High-Baseline-BMI and Increasing-BMI-Subgroups, with increased CPT-Commission/Omission-Errors (P-Group-Baseline-BMI=0.0001, P-Age-Interaction-Baseline-BMI=0.0046; P-Group-BMI-Trajectory=0.027, P-Age-Interaction-BMI-Trajectory=0.0729), and lower Hit-Reaction-Time (P-Group-Baseline-BMI=0.0001, P-Age-Interaction-Baseline-BMI=0.0046; P-Group-BMI-Trajectory=0.027, P-Age-Interaction-BMI-Trajectory=0.0729). See Figure-2-Panels-5A/5B and 6A/6B.

The High-Baseline-BMI subgroup also presented worsening attentional difficulties in the, reflected in progressively increasing CPT-Omission-Errors-T-Scores (P<0.0001) and Hit-Reaction-Time-T-Scores over time, along with a marginal reduction in Performance-IQ (P=0.047) driven by reduced Processing-Speed (P=0.012) (Supplementary-Figure-12).

### Chronic metabolic dysregulation predicts psychosis vulnerability mediating genetic vulnerability

The Increasing-BMI-Subgroup had significantly higher rates of Depressive Disorders (DD) (P=0.01), driven by a 30% prevalence in adolescent females, compared to a 13% adult lifetime prevalence in the Male-Decreasing-BMI-Subgroup (p=0.043, Chi-Square=8.13) (Figure-3-Panel-1A). The Increasing-BMI-Subgroup exhibited age-related worsening of depressive symptoms, including SIPS-Dysphoric-Mood (P-Group=0.0016, P-Age-Interaction=0.038), BPRS-Depression (P-Group=0.0003, P-Age-Interaction=0.005), BPRS-Guilt (P-Group=0.0013, P-Age-Interaction=0.057), particularly in the Female-Increasing-BMI subgroup. See Figure-3-Panels-1B-D.

The High-Baseline-BMI-Subgroup had an increased risk of developing a psychotic disorder (PD), with an 18% lifetime prevalence compared to 4% in the Low-Baseline-BMI-Subgroup (p=0.007, Chi-Square=7.09, HR=3.79, CI=1.37-10.5). Within the High-Baseline-BMI-Subgroup, PD risk was highest in males (25%) versus females (10%) See Figure-3-Panel-2A. Although all individuals were antipsychotic-naïve at baseline, the High-Baseline-BMI-Subgroup had a greater likelihood of receiving antipsychotic medication during follow-up up (P-Group-Effect=0.0005,P-Age-Interaction=0.0046), particularly in males (chi-square 8.9, p-value 0.01). See Supplementary-Figure-2.

**Figure 3:**
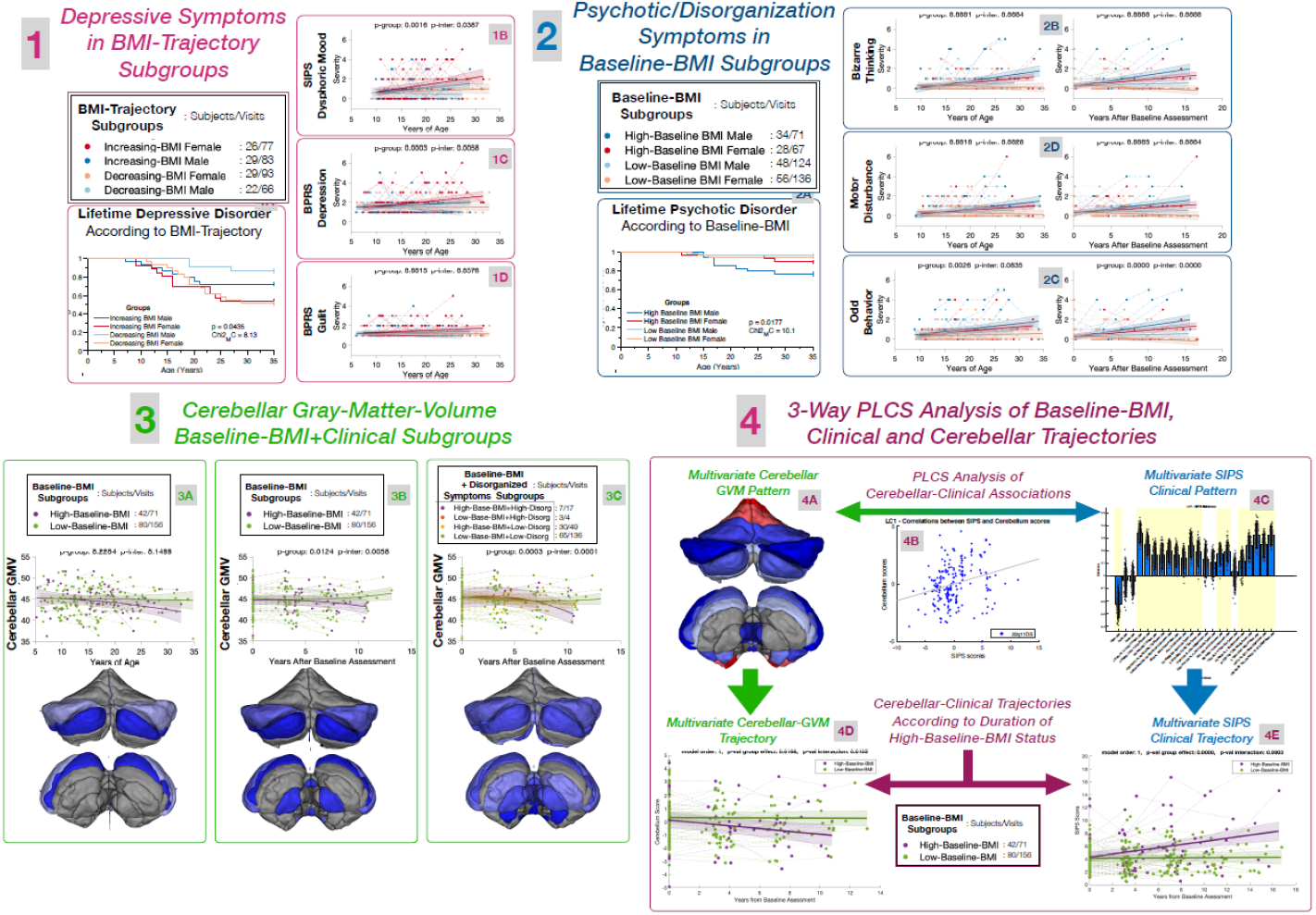
Downstream clinical and neurodevelopmental consequences of atypical BMI trajectories emerging during longitudinal follow-up. **Panels 1A-1D:** Differential vulnerability to depression during longitudinal follow-up in Increasing-vs-Decreasing-BMI-Trajectory Subgroups. Increasing-BMI-Males in dark blue, Increasing-BMI-Females in red, Decreasing-BMI-Males in light blue, and Decreasing-BMI-Females in orange. **Panel 1A:** Kaplan-Meyer survival curves for lifetime Depressive Disorder across BMI-Trajectory Subgroups. **Panel 1B:** Severity of the Dysphoric-Mood item of the SIPS compared across BMI-Trajectory Subgroups with Mixed-Model-Linear-Regression. **Panel 1C:** Severity of the Depression item of the BPRS compared across BMI-Trajectory Subgroups with Mixed-Model-Linear-Regression. **Panel 1D:** Severity of the Excessive Guilt item of the BPRS compared across BMI-Trajectory Subgroups with Mixed-Model-Linear-Regression. **Panels 2A-2D:** Differential vulnerability to psychotic disorders during longitudinal follow-up in High-vs-Low-Baseline-BMI-Subgroups. High-Baseline BMI-Males in dark blue, High-Baseline-BMI-Females in red, Low-Baseline-BMI-Males in light blue, and Low-Baseline-BMI-Females in orange. **Panel 2A:** Kaplan-Meyer survival curves for lifetime Psychotic Disorder across Baseline-BMI Subgroups. **Panel 2B:** Severity of the Bizarre-Thinking item of the SIPS compared across Baseline-BMI-Subgroups with Mixed-Model-Linear-Regression. In the left portion of the panel symptom severity is modelled according to age while in the right portion it is modelled according to time after baseline assessment. **Panel 2C:** Severity of the Motor-Disturbance item of the SIPS compared across Baseline-BMI-Subgroups with Mixed-Model-Linear-Regression. In the left portion of the panel symptom severity is modelled according to age while in the right portion it is modelled according to time after baseline assessment. **Panel 2D**: Severity of the Odd-Behavior item of the SIPS compared across Baseline-BMI-Subgroups with Mixed-Model-Linear-Regression. In the left portion of the panel symptom severity is modelled according to age while in the right portion it is modelled according to time after baseline assessment. **Panel 3A:** Trajectories of Cerebellar Gray Matter Volume (GMV) compared across baseline BMI-Subgroups using Mixed-Model-Linear-Regression, correcting for gender, psychotropic medication, intracranial volume, and scanner type. The upper panel displays the developmental trajectory of Total Cerebellar Gray Matter Volume. High-Baseline-BMI in Purple. Low-Baseline-BMI in Green. The anatomical pattern displays diverging developmental trajectories of 12 cerebellar subfields across subgroups. Subfields are color coded from dark to light blue according to p-value of age-interaction effect (darker blue indicating more significant differences). Subfields with non-significant differences after False Discovery Rate correction for multiple comparisons are displayed in gray. **Panel 3B:** Trajectories of Cerebellar Gray Matter Volume (GMV) compared across baseline BMI-Subgroups using Mixed-Model-Linear-Regression, correcting for gender, psychotropic medication, intracranial volume, and scanner type. Neurodevelopmental trajectories are modelled according to time from baseline assessment to characterize to investigate whether divergent cerebellar trajectories across Baseline-BMI subgroups were more directly predicted by duration of High-vs-Low BMI status that by the effect of age. **Panel 3C:** Trajectories of Cerebellar Gray Matter Volume (GMV) compared across baseline BMI-Subgroups and Clinical Subgroups using Mixed-Model-Linear-Regression, correcting for gender, psychotropic medication, intracranial volume, and scanner type. We modelled separate trajectories for Baseline-BMI subgroups divided according to whether or not they developed at least moderate (3 or more intensity on the SIPS Scale) motor, cognitive or behavioral disorganization symptoms, during longitudinal follow-up: High-Baseline-BMI/High-Symptoms in Purple, High-Baseline-BMI/Low-Symptoms in yellow, Low-Baseline-BMI/High-Symptoms in dark orange, and Low-Baseline BMI/Low-Symptoms in green. Cerebellar trajectories are modelled according to years after baseline assessment. This analysis investigated whether deviant cerebellar trajectories, developing as a function of duration of High-vs-Low BMI status were also linked to the development of disorganization symptoms during longitudinal follow-up. **Panel 4A:** Multivariate cerebellar pattern, derived from PLCS analysis capturing impact of multivariate SIPS clinical pattern on age-related Cerebellar-GMV trajectory. Color coding reflects loading of cerebellar lobules on the multivariate pattern, with Blue indicating significant positive loading and Red significant negative loading. **Panel 4B:** Association of Multivariate Cerebellar-GMV patterns and SIPS-Clinical patterns detected from PLCS analysis. **Panel 4C:** Multivariate SIPS clinical pattern, derived from PLCS analysis, capturing the impact of SIPS-Variables on age-related Cerebellar-GMV trajectories. The pattern detected a negative effect of age on Cerebellar-GMV. Most SIPS-Diagnosis-by-Age variables loaded positively onto this pattern, indicating that age-related Cerebellum-GMV reductions were steeper in individuals with higher symptom intensity. **Panel 4D:** Trajectories of Multivariate Cerebellar-GMV score derived from Cerebellar-Clinical PLCS analysis, according to Duration of High-vs-Low BMI status. **Panel 4E:** Trajectories of Multivariate SIPS-Clinical score derived from Cerebellar-Clinical PLCS analysis, according to Duration of High-vs-Low BMI status.

The High-Baseline-BMI-Subgroup also showed a progressive worsening of motor and thought disorganization symptoms, indicated by BPRS and SIPS Motor-Disturbances (P-Group=0.0016, P-Age-Interaction=0.0028) and BPRS Bizarre-Behavior (P-Group=0.0001, P-Age-Interaction=0.0004) (Figure-3-Panels-2B-C-Left). Additional disorganization symptoms, including SIPS-Odd-Behavior (P-Group=0.0026, P-Age-Interaction=0.083), SIPS/BPRS-Self-Neglect, and SIPS-Disorganized-Communication, were elevated but showed no significant difference in age-related developmental trajectory. See Supplementary-Figures-9/10.

Interestingly, disorganization symptom trajectories diverged significantly across Baseline-BMI-Subgroups when modeled according to time from baseline assessment. In the High-Baseline-BMI group, symptoms such as SIPS-Odd-Behavior (P-Group<0.0001, P-Time-from-Baseline-Interaction<0.0001) progressively worsened, with a stronger link to High-BMI duration rather than age alone (Figure-3-Panels-2D-Right. Supplementary-Figures-13/14).

### Cerebellar maturation links prolonged High-BMI status to psychosis vulnerability

BMI-Trajectory subgroups presented similar Cortical and Cerebellar GMV developmental trajectories. See Supplementary-Figure-17-18. Baseline-BMI subgroups did not significantly differ in terms of Cortical GMV trajectories. See Supplementary-Figure-18. We instead observed a significant age-related reduction of Cerebellar-GMV in the High-Baseline-BMI-Subgroup, that specifically affected the posterior/inferior lobules and was not significant for total Cerebellar-GMV (P-Group=0.22, P-Age-interaction=0.149). See Figure-3-Panel-3A, Supplementary-Figure-17. The trajectory of Total-Cerebellar-GMV however significantly differed across Baseline-BMI-subgroups when it was modelled according to Time-From-Baseline-Assessment (P-Group-Effect=0.012, P-Time-from-Baseline-Interaction=0.005), with progressive Total-Cerebellar-GMV reductions developing as a function of duration of High-BMI-Status and driven by the posterior-inferior lobules. See Figure-3-Panel-3B. Modelling the joint effects of Baseline-BMI and disorganization symptoms revealed an even broader pattern of Cerebellar-GMV reductions, predicted by duration of High-BMI-Status and driven by High-Baseline-BMI individuals who developed significant disorganization symptoms during longitudinal follow-up (P-Group-Effect=0.0003, P-Time-from-Baseline-interaction=0.0001). See Figure-3-Panel-3C. These results suggest that Cerebellar-GMV reductions associated with High-Baseline-BMI were more directly predicted by a duration-dependent effect of prolonged High-BMI status, than by other age-related factors, and were furthermore linked to the development of disorganization symptoms.

We then investigated whether Baseline-BMI status mediated the association between multivariate clinical and neurodevelopmental trajectories. PLCS analysis identified a single significant component accounting for the effect of SIPS-intensity diagnosis on the developmental trajectory of Cerebellum-GMV (r = 0.27, p = 0.002; see Figure-3-Panel-4). Specifically, PLCS detected a negative effect of age on overall Cerebellum-GMV, primarily driven by reductions in the posterior-inferior cerebellar lobules. See Figure-3-Panel-4A. Most SIPS-Diagnosis-by-Age variables loaded positively onto this pattern, indicating that age-related Cerebellum-GMV reductions were steeper in individuals with higher symptom intensity. See Figure-3-Panel-4C. SIPS variables that most significantly differentiated the Cerebellum-GMV trajectory included Bizarre Thinking, Odd Behavior, Disorganized Communication, Impaired Personal Hygiene, and Unusual Thought Content. See Supplementary-Figure-20. From this PLCS analysis, we derived a multivariate SIPS-Clinical Score and a Cerebellum-GMV Score by matrix-multiplying the loadings of SIPS and Cerebellum-GMV variables with values measured at each longitudinal visit. Next, we tested whether BMI status differentially modulated the developmental trajectories of these multivariate scores. Results showed that High-Baseline-BMI contributed to both a sharper age-related increase in the SIPS-Clinical Score (P-Group=0.0026, P-Age-interaction=0.02) and sharper age-related decline in the Cerebellum-GMV Score (P-Group=0.042, P-Age-interaction=0.034). See Supplementary-Figure-19. The divergence in both the SIPS-Clinical Score (P-Group-Effect<0.0001, P-Time-from-Baseline-interaction<0.0001) and Cerebellum-GMV Score (P-Group-Effect=0.016, P-Time-from-Baseline-interaction=0.01) was even more pronounced when modeling the effect of time from the baseline assessment. See Figure-3-Panel-4D-E. This would suggest that High-BMI status may mediate the association between increasing SIPS intensity and the decline in posterior-inferior Cerebellum-GMV, through a dose-effect relationship with High-BMI status duration.

We then performed the same PLCS analysis to examine the link between SIPS-intensity diagnosis and Cortical-GMV values. Results (detailed in the Supplementary-Figures 21/22) identified three significant components capturing the differential effects of SIPS clinical patterns on Cortical-GMV trajectories. While High-Baseline-BMI status differentially moderated the clinical trajectory of SIPS-Clinical Scores derived from all three components, it did not significantly affect the developmental trajectory of Cortical-GMV Scores. This suggests that the clinical correlates of High-Baseline-BMI status may be specifically related to its effects on posterior-inferior Cerebellum GMV reductions, whereas Baseline-BMI status did not significantly mediate the association between clinical and Cortical-GMV trajectories.

## Discussion

We explored the developmental impact of metabolic dysregulation on psychosis vulnerability in a large longitudinal cohort of individuals with 22q11DS. The behavioral and neurocognitive impulsivity dimension of ADHD during childhood was associated with subsequent chronic metabolic dysregulation. Childhood metabolic dysregulation contributed to a progressive increase in psychosis vulnerability. A dose-effect relationship linked prolonged metabolic dysregulation to progressive alterations in cerebellar morphology, which were related to the development of motor and disorganization symptoms of psychosis. These results identify a novel neurodevelopmental-metabolic pathway linking childhood inhibitory control to psychosis vulnerability through cerebellar dysmaturation, described schematically in Figure 4.

**Figure 4:**
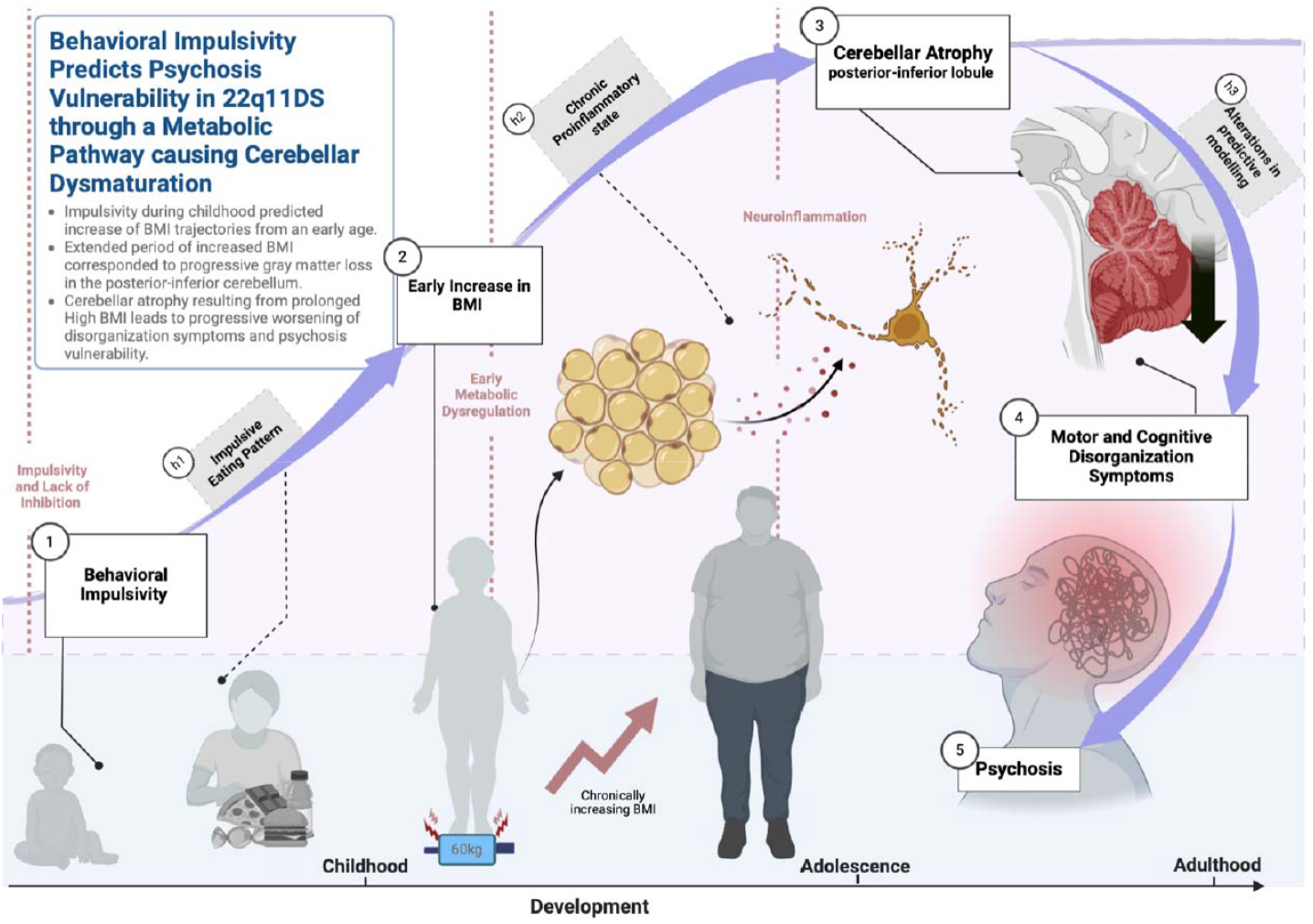
Schematic representation of a possible developmental pathways accounting for the main results and discussed above. **White boxes** highlight correlates of atypical BMI trajectories observed at different developmental stages. **Box 1:** Behavioral impulsivity evident particularly during childhood predicts subsequent alterations in BMI Trajectory. **Box 2:** Deviations in BMI trajectories in 22q11DS emerge during childhood and progressively worsening during adolescence and early adulthood, preceding the prescription of antipsychotic medication. **Box 3:** Atrophy of the Posterior Inferior Cerebellum develops progressively as a result of chronic metabolic dysregulation following a dose-effect relationship with duration of High-BMI Status. **Box 4:** Progressive cerebellar atrophy is linked to the development of motor and cognitive disorganization symptoms that also follow a dose effect relationship with duration of High-BMI status. **Box 5:** Progressive worsening of disorganization symptoms results in increased vulnerability of being diagnosed with a psychotic disorder requiring antipsychotic treatment as a result of chronic metabolic dysregulation. **Gray boxes** highlight non-measured mechanisms that could hypothetically link the different developmental alterations. **Box H1:** Impulsive eating patterns could mediate the link between behavioral impulsivity and early chronic BMI increases reflecting shared impairment of cortico-striatal circuits responsible for top-down inhibitory control goal-directed and feeding behavior. **Box H2**: Chronic pro-inflammatory stated linked to prolonged metabolic dysregulation could affect cerebellar maturation and contribute to account for the dose effect relationship linking duration of metabolic dysregulation to progressive cerebellar atrophy. **Box H3:** Alterations in the posterior inferior cerebellum could impair predictive modelling of higher order cognitive processes resulting in motor and cognitive disorganization symptoms of psychosis and progressive worsening of executive and attentional difficulties.

### Behavioral impulsivity predicts BMI trajectories

Recent studies in the general population suggest a significant overlap in the genetic underpinnings of ADHD with both obesity (11) and psychosis (35) but the pathophysiological mechanisms accounting for these associations remain largely unclear. Our results show for the first time that in 22q11DS genetic vulnerability to chronic metabolic dysregulation is linked to the presence of behavioral and neurocognitive impulsivity during childhood. Indeed, both early BMI-increases and progressive increases in BMI scores were associated with diagnoses of impulsive ADHD, higher childhood externalizing psychopathology and neurocognitive trajectories indicative of childhood impulsivity. While these results are novel in 22q11DS, they are in line with evidence in the general population indicating that impulsivity/hyperactivity dimension of ADHD strongly predicts the emergence of early-life obesity (15). It has been proposed that the impulsivity dimension of ADHD may contribute to impulsive eating patterns (17), which might reflect overleaping alterations in cortico-striatal circuits underlying top-down inhibitory control of feeding and goal directed behavior (36). Our results suggest that the deleterious psychological and neurodevelopmental consequences of early-life obesity may contribute to the genetic and clinical link between childhood ADHD and adult psychopathology (18).

### Chronic metabolic dysregulation predicts psychosis vulnerability mediating genetic vulnerability

Recent epidemiological evidence linked early deviations in metabolic trajectories to subsequent psychosis vulnerability (4). However, behavioral phenotyping was only performed at the end of longitudinal follow-up, limiting the ability to characterize behavioral precursors of metabolic dysregulation (4). The unique 22q11DS model allowed us to characterize both the metabolic and clinical trajectories preceding the emergence of psychosis. Metabolic dysregulation preceded and predicted the subsequent transition from prodromal symptoms to full-blown psychotic disorders, which was more strongly related to the duration of high-BMI status than other age-related factors. These findings strongly argue against reverse causal mechanisms and would rather suggest a dose effect-relationship linking duration of metabolic dysregulation with subsequent risk of psychosis (2, 4). This metabolic pathway significantly mediated the effects of genetic vulnerability to psychosis in 22q11DS.

Converging evidence suggests that the prolonged immunological dysregulation associated with obesity could contribute increased vulnerability to psychosis (2, 4). Firstly, in the above-mentioned epidemiological study, the effects of increased BMI on psychosis vulnerability were mediated by the development of insulin resistance and increase in pro-inflammatory markers(4). Secondly, increased pro-inflammatory markers have been consistently reported at all stages of psychosis vulnerability(2), predicting the subsequent emergence of psychosis (4, 37) and partially explaining shared genetic and environmental risk among relatives (37). Moreover, immunological factors could directly alter adolescent synaptic pruning implicated in the pathophysiology of psychosis (24). However, the underlying mechanisms contributing to the development of immunological dysregulation in psychosis have remained unclear(2, 22). Interestingly, obesity is possibly the strongest risk factor for the development of chronic pro-inflammatory states (23). Moreover, childhood obesity has a strong tendency to persist through adolescence and into adulthood (38), implying that obese children are potentially exposed to a chronic pro-inflammatory state throughout the critical vulnerability period of adolescent brain development (4). Indeed, the three-way association between obesity, immunological dysregulation and psychosis, is increasingly documented in the general population (4) and has been recently observed in 22q11DS (39).

Our findings suggest a dose-effect relationship between duration of metabolic dysregulation and subsequent vulnerability to psychosis, which is compatible with a progressive effect of prolonged immunological dysregulation on neurodevelopmental processes associated with psychosis. Our clinical results suggest that brain structures implicated in motor and cognitive disorganization symptoms of psychosis might be particularly sensitive to the effects of metabolic dysregulation.

### Cerebellar maturation links chronic metabolic dysregulation to psychosis vulnerability

While cortical GMV trajectories were not significantly affected by BMI status, we observed a significant reduction in cerebellar GMV in individuals with high baseline BMI. Moreover, cerebellar GMV reductions were more tightly related to the duration of high BMI status than to age, suggesting a potential dose-response relationship linking prolonged obesity to posterior-inferior cerebellar GMV atrophy. Reductions of posterior-inferior cerebellar GMV are among the strongest neuroanatomical correlates of both psychosis (20) and obesity (19). Moreover, cerebellar alterations observed in first-episode psychosis patients, might be directly explained by co-occurring metabolic (25) and immunological dysregulation (40). Finally, cerebellar dysfunction could mechanistically contribute to key hallmarks of psychosis symptomatology, including impaired adaptation of predictive models of the world based on past experience (41), leading to disorganized thought and behavior (42).

We investigated whether posterior-inferior cerebellar GMV atrophy was linked to developmental increases in motor and cognitive disorganization symptoms associated with prolonged high BMI status. Our findings demonstrated that duration-dependent cerebellar GMV atrophy was particularly severe in individuals with high baseline BMI who also developed disorganization symptoms during longitudinal follow-up. Furthermore, multivariate analysis—blind to BMI status—revealed that posterior-inferior cerebellar GMV atrophy was specifically linked to more severe psychosis symptoms, particularly disorganization symptoms. Notably, clinical and neurodevelopmental trajectories reflecting such association diverged progressively based on the duration of high vs. low baseline BMI status. This suggests that a dose-response relationship with prolonged High-BMI status may directly contribute to the developmental association between posterior-inferior cerebellar atrophy and the emergence of disorganization symptoms. Interestingly, while we also detected associations between SIPS clinical patterns and cortical GMV trajectories, these were not significantly influenced by BMI status.

Overall, these findings suggest that cerebellar sensitivity to metabolic dysregulation may contribute to the characteristic clinical trajectory associated with prolonged high BMI status. Although further investigation is warranted, emerging translational evidence suggests that the cerebellum may play a key role in mediating immunological responses to systemic infection through increased blood-brain barrier permeability compared to the cerebral cortex (43). The 22q11DS syndrome is associated with disruptions in blood-brain barrier permeability (44), which could further heighten cerebellar vulnerability to the negative immunological effects of prolonged metabolic dysregulation. This immunological-cerebellar pathway may contribute to the emergence of motor, cognitive, and behavioral disorganization symptoms—hallmarks of psychotic disorders—following chronic metabolic dysregulation. Additionally, the posterior-inferior cerebellum is directly implicated in attentional and executive functioning. (45). Thus, cerebellar alterations may significantly contribute to the progressive worsening of attentional difficulties associated with high baseline BMI. This metabolic-cerebellar pathway could link early behavioral impulsivity to the persistence and worsening of attentional and executive dysfunction—patterns commonly observed in ADHD (46).

### Strengths and Limitations

The main strength of the present study pertains to the long-term multimodal longitudinal phenotyping of 22q11DS, which provided a unique opportunity to characterize the developmental pathways linking genetic vulnerability to metabolic dysregulation, ADHD and psychosis, from childhood to adulthood. Still, the present results should be considered in the light of some limitations. Firstly, the temporal directionality linking early metabolic dysregulation and subsequent psychosis vulnerability is not inherent proof of causality. However, neurodevelopmental and clinical alterations were more tightly related to duration of metabolic dysregulation, than to other age-related factors, which support a dose-effect causal relationship. Secondly, we did not have access to biochemical measures of immune dysregulation, which according to our model, should significantly mediate the effects of BMI on clinical and neurodevelopmental trajectories. However, a strong three-way association between BMI, immune dysregulation and psychosis has been reported in an independent cohort of individuals with 22q11DS (39), which would support the candidate role of immune dysregulation in our study. Thirdly, as detailed below, the generalizability of our findings to the general population warrants further investigation.

### Conclusions and future directions

Our results suggest that chronic childhood metabolic dysregulation could significantly mediate the effects of genetic vulnerability to psychosis in 22q11DS, by affecting cerebellar maturation. These results support a causal link underlying the association of metabolic dysregulation with psychosis vulnerability described in the general population (4), warranting further confirmation by future studies. Firstly, it would be important to further clarify the mechanisms leading to metabolic dysregulation in 22q11DS. Our results suggest that alterations in top-down inhibitory control underlying childhood behavioral impulsivity might lead to impulsive eating patterns that contribute to chronic metabolic dysregulation. Secondly, additional mechanisms might increase vulnerability to the effects of early life obesity in 22q11DS, such as increased susceptibility to immunological consequences of obesity(23), or increased neurodevelopmental vulnerability to chronic pro-inflammatory states, due to increased blood-brain-barrier permeability (44). Thirdly, future studies could explore mechanisms linking pubertal BMI increases and female-predominant vulnerability to depressive symptoms, which also strikingly recapitulates findings in the general population (4). For instance, early-life obesity might alter Hypothalamus-Pituitary-Adrenal-Axis maturation, which has been linked to the development of anxio-depressive symptomatology in 22q11DS (26). Fourthly, our findings highlight the importance of further clarifying the impact of metabolic dysregulation on the clinical trajectory of ADHD beyond 22q11DS, including its role in mediating the association with psychosis vulnerability (18, 35). From a clinical perspective, our results suggest that targeting the metabolic consequences of behavioral impulsivity could reduce psychosis vulnerability and improve neurocognitive trajectories in 22q11DS, as well as potentially in the general population. As such, our results provide a strong rationale for trials assessing the role of early interventions tackling early-life metabolic dysregulation and obesity as a strategy to reduce the risk of psychosis.

## Data Availability

All data produced in the present study are available upon reasonable request to the authors

## Acknowledgments

This study was supported by the Swiss National Science Foundation (SNSF) (Grant numbers: to SE FNS 320030_179404, FNS 324730_144260) and by the National Center of Competence in Research (NCCR) Synapsy-The Synaptic Bases of Mental Diseases (Grant number: 51AU40_125759). Prof Maude Schneider (#162006) and Dr Corrado Sandini (#209096) were supported by grants from the SNF. Samuele Cortese, NIHR Research Professor (NIHR303122) is funded by the NIHR for this research project. The views expressed in this publication are those of the author(s) and not necessarily those of the NIHR, NHS or the UK Department of Health and Social Care. Samuele Cortese is also supported by NIHR grants NIHR203684, NIHR203035, NIHR130077, NIHR128472, RP-PG-0618-20003 and by grant 101095568-HORIZONHLTH-2022-DISEASE-07-03 from the European Research Executive Agency. Figure-4 is created with Biorender.com. We warmly thank all the families who participated in the study.

