## Supplementary material for "Chronic early-life obesity linked to childhood impulsivity predicts long-term psychosis trajectory through dose-dependent cerebellar dysmaturation in 22q11.2 Deletion Syndrome"

**Table of Contents**

1.	Methods	2

1.1	Participants:	2

1.2	Clinical Instruments Employed for Psychiatric Assessment:	2

1.3	Statistical Analyses	2

1.4	Structural MRI Image Processing and Analysis	3

2.	Results	5

2.1	Comparison of demographic characteristics of across 22q11DS and Healthy Controls and across 22q11DS BMI subgroups	5

2.2	BMI Trajectories compared across 22q11DS and Healthy Controls considering only assessments when individuals were still naïve to antipsychotic medication.	5

2.3	Comparison of Psychotropic Medication across BMI-Subgroups	6

2.4	Comparison of DSM-5 Psychiatric Diagnoses across BMI Subgroups	8

2.5	Comparison of Trajectories of individual Clinical/Neurocognitive Scores across BMI Subgroups	12

2.6	Comparison of Trajectories of Gray Matter Volume (GMV) across BMI Subgroups	16

2.7	PLCS Analysis of 3-way Clinical-Cerebellar-GMV-BMI Association	19

2.8	PLCS Analysis of 3-way Clinical-Cortical-GMV-BMI Association	22

**Figures**

**Tables**

### 1. Methods

#### 1.1 Participants:

Both healthy controls (HCs) and individuals with 22q11DS were recruited in the context of an ongoing longitudinal study that has been described extensively in previous publications (1, 2). Participants with 22q11DS were recruited through patient associations or word-of-mouth throughout French and English-speaking European countries. HCs were mostly recruited among non-affected siblings. All participants and parents provided written informed consent under protocols approved by Ethics Committee Geneva. Once recruited in the study participants were then followed up longitudinally approximately every 3 years. For the present study, we recruited 372 participants, including 184 participants with 22q11DS (M/F=93:91) and 188 HCs M/F=92:96), how were followed up for a total of 765 longitudinal assessments (433 in 22q11DS and 333 in HCs). Groups did not differ in terms of age at baseline assessment (13.2(6.6) in 22q11DS, 12.9(5.8 in HCs, p=0.66). However, number of longitudinal follow-up assessments were significantly higher in 22q11DS samples (2.3(1.2) in 22q11DS, 1.7(0.85) in HCs, p<0.001), leading to longer average duration of longitudinal follow-up (5.1(4.7) in 22q11DS, 2.9(3.3) in HCs, p<0.001). See Table 1 in the main text for a full description of demographic features.

#### 1.2 Clinical Instruments Employed for Psychiatric Assessment:

Psychiatric diagnoses were assessed according to the DSM-V (APA, 2013). We used the diagnostic Interview for Children and Adolescents-Revised and the psychosis supplement from the Kiddie Schedule for Affective Disorders and Schizophrenia for individuals below 18 years of age (3, 4), and the Structured Clinical Interview for DSM-V Axis I Disorders (5) for adults. These were complemented with the Structured Interview for Prodromal Syndromes (SIPS) (6), the Brief Psychiatric Rating Scale (BPRS) (7) and the Child/Adult Behavior Checklist (CBCL/ABCL) (8, 9). Intelligent quotient (IQ) was measured using the Wechsler Intelligence Scale for Children (WISC-III or IV)(10) and Wechsler Adult Intelligence Scale (WAIS-III or IV) (11)). Finally, age-normed T-Scores of Omission and Commission errors at the Conners’ Continuous Performance Test 2nd edition, CPT2 were used to measure inattention and impulsivity (12).

#### 1.3 Statistical Analyses

- *Mixed-Models-Linear Regression analysis of developmental trajectories.*

We employed mixed-models linear regression (MMLR), detailed in previous publications, implemented through a previously published toolbox: <https://github.com/danizoeller/myMixedModelsTrajectories> (1, 13), to characterize and test differences in developmental trajectories of BMI scores, firstly between HCs and 22q11DS (14, 15), modelling for the effects of gender. Briefly, models of increasing order, from constant to cubic, were fit to each variable being tested and a Bayesian information criterion (BIC)-based model selection method was employed to select the optimal model order. We employed a likelihood ratio test to evaluate differences in trajectories between groups both in terms of age-interaction effects (curves that do not follow a parallel developmental path) and intercept differences (curves that follow parallel developmental paths at different intercepts). As confirmatory analysis we repeated the analysis including only time-points in which 22q11DS had not been previously or currently treated with antipsychotic medication.

- *Subgroup analysis of BMI trajectories.*

We then identified subgroups of 22q11DS who presented diverging BMI trajectories, in order to explore clinical and neurodevelopmental correlates, as described elsewhere(16). Specifically, we firstly defined two normative BMI trajectories for male and female 22q11DS individuals using MMLR. We then separated 22q11DS individuals in two distinct subgroups according to whether their BMI score at the first baseline assessment was higher or lower than the age-equivalent gender-specific normative trajectory, which yielded two High-Baseline BMI and Low-Baseline-BMI subgroups. In a second complementary analysis we separated individuals according to whether the longitudinal rate of change of BMI was higher or lower than what was predicted by the age-equivalent gender-specific normative trajectory, yielding Increasing-BMI and Decreasing-BMI subgroups. To minimize the potential reversal-causal effects of antipsychotic medication on metabolic trajectories, we defined normative BMI trajectories, and assigned individuals to different BMI-subgroups, considering only assessments when participants were still naïve to antipsychotic medication. Subgroups did not significantly differ in any demographic measure, as described in Table 1 of the main text.

- *Statistical Comparison of Discrete Outcomes*

BMI-subgroups we also compared in terms of proportion of discrete outcomes, including the use of Antipsychotics (AP), Selective Serotonin Reuptake Inhibitors (SSRIs) and Psychostimulants (PST) medication, and the presence of DSM-V diagnosis of psychotic disorders, mood disorders, anxiety disorders, and ADHD diagnoses that were separated in purely Inattentive-ADD and Combined-ADHD subtypes, according to the presence of significant Impulsivity/Hyperactivity traits, defined as >3 Impulsivity/Hyperactivity symptoms. We firstly constructed Kaplan-Meyer survival curves by measuring the lifetime prevalence of discrete outcomes across subgroups, considering assessments performed at increasing ages, in successive 1-year iterations which we compared across BMI-subgroups, using log rank test, implemented in Matlab (<https://github.com/aebergl/MatSurv>), both considering separate and merged gender subgroups. We also measured the prevalence of discrete outcomes that persisted until the end of longitudinal follow-up, which we compared across BMI-subgroups using a 2-sample chi-square test, when ignoring the effect of gender, and a 4-sample chi-square test, when considering gender subgroups separately.

#### 1.4 Structural MRI Image Processing and Analysis

- *CERES Approach to Cerebellar Segmentation*

Cerebellar segmentation was achieved using the CERES method implemented on the online Volbrain MRI volumetry platform (17). The CERES algorithm is based on a multiatlas label fusion technique(18), trained against expert manual segmentation of the cerebellum in a collection of T1-weighted MRI scans. The outlining of all structures follows the guidelines and definitions summarized in the study by (19). The quality of cerebellar segmentation was manually checked with visual inspection by a single investigator (LP). For the present study we explored cerebellar trajectories only within the 22q11DS sample, is association with BMI and clinical trajectories. 3T-T1-weighted images, with high-quality cerebellar segmentation were available for a total of 139 22q11DS individuals and 254 assessments, from which we extracted measures of Gray-Matter-Volume (GMV) at the level of the entire cerebellum and of 12 cerebellar lobules, averaged across bilateral cerebellar hemispheres. Cerebellar GMV measures were corrected for gender, scan-type, psychotropic medication and intracranial volume with linear regression.

- *Freesurfer Approach to Cortical Segmentation*

T1-weighted structural MRI data were processed using FreeSurfer v7.4.1 The pipeline included intensity normalization, skull stripping, cortical surface reconstruction, and anatomical parcellation. Longitudinal T1-weighted scans were available for 139 participants with 22q11DS, yielding a total of 254 time points. Cortical gray matter volumes were extracted from 34 regions of interest (ROIs) per hemisphere based on the Desikan-Killiany atlas (20). Corresponding ROIs from the left and right hemispheres were combined and subsequently adjusted for sex, scan type, psychotropic medication use, and intracranial volume using linear regression

- *Subgroup analysis of Cerebellar Trajectories based on Baseline BMI and clinical trajectories.*

In order to test the presence of a 3-way interaction between duration of increased BMI status and subsequent clinical and neurodevelopmental outcomes, we defined subgroups according to both Baseline-BMI and the presence of SIPS symptoms of Motor-Disturbances, Bizarre-Thinking and Odd-Behavior a least moderate intensity defined as a score of at least 3/6 on the SIPS scale. These disorganization symptoms were selected due to their strong association with prolonged high-BMI status. We then compared neurodevelopmental trajectories across the 4 resulting subgroups (High-Baseline-BMI/High-Symptoms, High-Baseline-BMI/Low-Symptoms, Low-Baseline-BMI/High-Symptoms, Low-Baseline-BMI/Low-Symptoms) described in Supplementary Table 1. Demographic features described in Supplementary Table 1 refer to subjects/assessments from whom high quality MRI segmentation of Cerebellar and Cortical Gray-Matter Volume was available and that were analyzed for the comparison of neurodevelopmental trajectories, described in the main text. By comparing neurodevelopmental trajectories across subgroups defined according to both Baseline-BMI and development of disorganization symptoms we tested whether progressive cerebellar atrophy linked to duration of High-BMI status, was also linked to the development of disorganization symptoms during longitudinal follow-up.

- *Partial Least Square Correlation Analysis of Multivariate Clinical-Neurodevelopmental Association Patterns.*

We employed multivariate Partial Least Square Correlation Analysis (PLCS) to identify significant associations between clinical symptoms and neurodevelopmental trajectories, as well as to assess whether BMI status contributed to these associations. PLCS uses singular value decomposition to reveal relationships between data types, which are represented as bar plots. The height and direction of the bars indicate the contribution of individual variables (positive vs. negative). Statistical significance was assessed using permutation testing, while bootstrapping identified variables that consistently contributed to the patterns (highlighted in yellow).

We conducted two separate PLCS analyses to investigate the clinical correlates of cerebellar and cortical gray matter volume (GMV) trajectories. This method has been extensively used to examine whether specific clinical diagnoses, such as the presence of significant positive symptoms, are associated with deviations in multivariate neurodevelopmental trajectories by moderating the effect of age on neurobiological measures (21, 22). In this study, rather than predefining clinical correlates, we differentiated individuals based on whether they developed symptoms of at least moderate intensity (≥3/6) for each of the 19 symptoms assessed using the Structured Interview for Prodromal Syndromes (SIPS), resulting in 19 distinct diagnostic vectors.

We constructed a behavioral matrix comprising age and 19 age-by-diagnosis vectors, which was then contrasted against two neurodevelopmental matrices. These matrices contained either 12 cerebellar GMV measures or 34 cortical GMV measures. This analysis detected multivariate clinical patterns that differentially moderated deviations in neurodevelopmental trajectories.

When significant brain-behavior correlations were identified through PLCS analysis, we extracted multivariate behavioral and brain scores by matrix multiplication of variable loadings with the clinical and GMV values measured at each longitudinal assessment. Finally, we evaluated whether the developmental trajectories of these multivariate behavior/brain scores differed significantly based on BMI status. This would indicate that BMI status significantly contributed to the brain-behavior correlations detected through PLCS analysis.

### 2. Results

##### 2.1 Comparison of demographic characteristics of across 22q11DS and Healthy Controls and across 22q11DS BMI subgroups

22q11DS and Healthy Controls did not differ in terms of age at baseline assessment (13.2(6.6) in 22q11DS, 12.9(5.8 in HCs, p=0.66). However, number of longitudinal follow-up assessments were significantly higher in 22q11DS samples (2.3(1.2) in 22q11DS, 1.7(0.85) in HCs, p<0.001), leading to longer average duration of longitudinal follow-up (5.1(4.7) in 22q11DS, 2.9(3.3) in HCs, p<0.001). This difference is likely related to the difficulty in motivating healthy controls to engage in time-consuming longitudinal assessments, in the absence of strong motivation for high quality clinical assessment provided to 22q11DS individuals. Several considerations make us confident that such demographic differences do not impact the results and conclusions detailed in the main text. Firstly, the association between 22q11DS and atypical BMI trajectories consistent with our results are extensively documented in previous literature (23, 24)making it highly unlikely that demographic differences are responsible for BMI trajectory alterations observed in our data. Secondly, the core findings described in the main text relate to the clinical and neurodevelopmental correlates of diverging BMI trajectories within subgroups of 22q11DS individuals. Importantly, neither BMI-Trajectory nor Baseline-BMI subgroups presented significant differences in demographic features, including duration of longitudinal follow, which therefore is unlikely to significantly influence BMI trajectory alterations described in 22q11DS and their clinical and neurodevelopmental correlates.

##### 2.2 BMI Trajectories compared across 22q11DS and Healthy Controls considering only assessments when individuals were still naïve to antipsychotic medication.

To verify that prescription of Antipsychotic medication was not responsible for atypical BMI trajectories observed in 22q11DS we repeated the comparison of BMI trajectories considering only assessments when individuals were still naïve to antipsychotic medication (66 Assessments in 38 Subjects) resulting in a total of 364/433 assessments 166/184 individuals, which we compared with BMI trajectories in healthy controls. We observed striking differences in BMI trajectories in Antipsychotic-Naïve 22q11DS compared to Healthy Controls, which were characterized by a steeper increase in BMI scores during childhood and early adolescence (p-interaction <0.001) leading to significantly higher BMI scores by late-adolescence/early-adulthood (p-group-effect<0.001). See Supplementary Figure 1, Panel A. To appreciate the developmental link between BMI trajectory alterations and antipsychotic prescription we juxtapose the BMI trajectories without and with assessments in which 22q11DS individuals were prescribed antipsychotic medication identified by black asterisks in Supplementary Figure 1, Panel B. Comparing the Panels A and B of revealed that BMI increases in antipsychotic naïve 22q11DS individuals preceded the prescription of Antipsychotic medication. Prescription of antipsychotic medication was associated with a further acceleration of BMI increase during late-adolescence/early adulthood but was not responsible for BMI increases during childhood/early adolescence. In a further analysis described in the following results section we specifically investigated whether early BMI increases during childhood might have contributed increased risk of receiving antipsychotic medication during longitudinally follow-up, partially accounting for the association between antipsychotic medication and increased BMI.

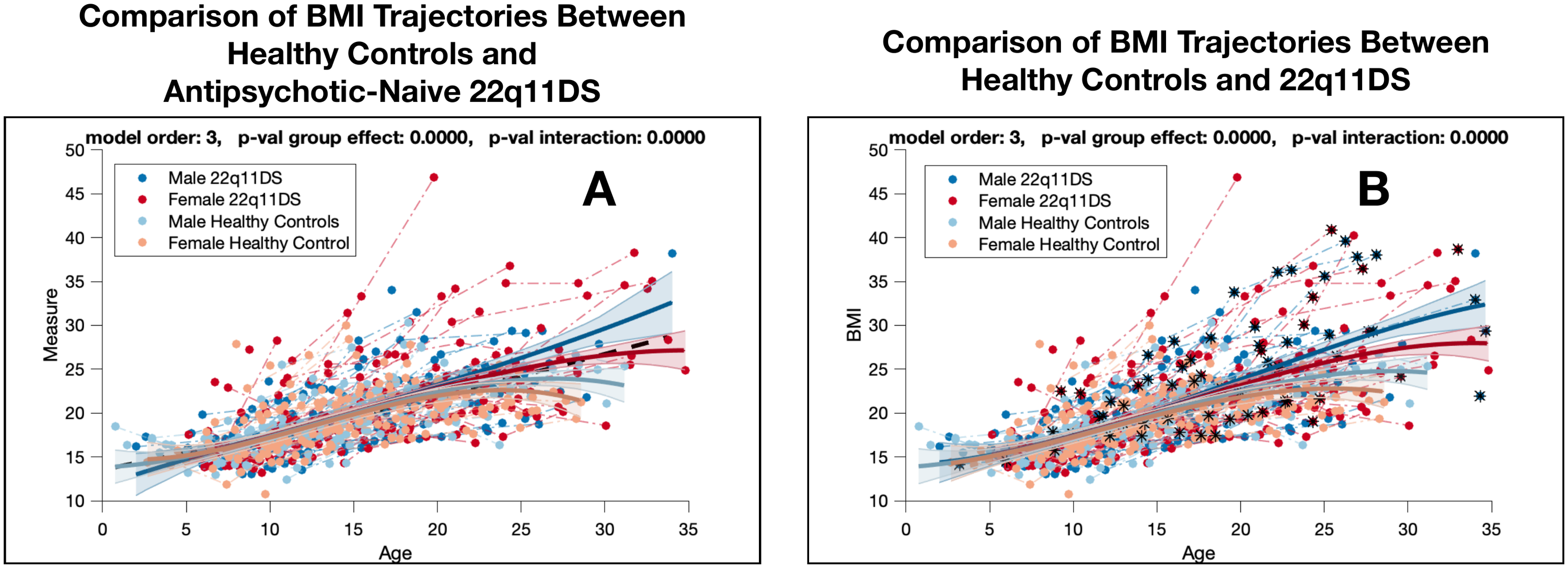

Supplementary Figure 1: Comparison of BMI Trajectories between HCs and 22q11DS

**Panel A**: Developmental trajectories of BMI scores compared across Healthy Controls and antipsychotic-naïve 22q11DS individuals modelling for the effects of gender yielding 4 subgroups: Male 22q11DS in dark blue, Female 22q11DS in red, Male Healthy Controls in Light Blue, Female Healthy Controls in Orange. **Panel B**: Developmental trajectories of BMI scores compared across Healthy Controls and 22q11DS also considering assessments during which 22q11DS individuals are receiving antipsychotic treatment, highlighted in black.

##### 2.3 Comparison of Psychotropic Medication across BMI-Subgroups

BMI-Subgroups had similar use of psychotropic medication with the exception of an association between Baseline-BMI and antipsychotic medication. The High-Baseline-BMI subgroup, who by design was naive two antipsychotic medication at baseline assessment had a higher likelihood of being prescribed antipsychotics during longitudinal follow up. This was captured by a significant age-interaction effect (p-age-interaction=0.0046), when modeling the trajectory of antipsychotic prescription at individual assessments using MMLR. See Panel A1 of Supplementary Figure 2. This difference was only partially explained by higher rates of lifetime antipsychotic prescription in the High-Baseline-BMI subgroup, as revealed by non-significant Kaplan-Meyer survival curve analysis described in Panels B1 and C1 of Supplementary Figure 1. However, in the High-Baseline-BMI had a higher likelihood of being prescribed chronic antipsychotic medication persisting until the end of longitudinal follow-up, particularly in the Male-High-Baseline-BMI subgroup (chi-square 8.9, p-value 0.01). See Panel D1 of Supplementary Figure 2.

Given that BMI subgroups were defined prior to antipsychotic prescription, these results would suggest that the association between increased BMI and AP medication is at least partially explained by an increased risk of being prescribed chronic antipsychotic medication in individuals with high baseline BMI. Considering the clinical results described in the main text, it is plausible that prolonged antipsychotic prescription may have resulted from the increased probability of developing a psychotic disorder during longitudinal follow up observed in the High-Baseline-BMI. We however cannot exclude that antipsychotic prescription might have contributed to a further and progressive increase in BMI scores observed in the High-Baseline-BMI subgroup.

Moreover, given that BMI-trajectory subgroups were also defined considering only antipsychotic naive individuals, it is plausible that a significant proportion of more highly symptomatic individuals requiring antipsychotic medication might have been excluded from this analysis. This conservative approach, hence potentially underestimated the clinical impact of longitudinal BMI increase, but on the other hand ensured that the clinical correlates of BMI trajectories were not related to the reversal causal effect of antipsychotic medication.

It should also be noted that limited information was available concerning the compliance to psychotropic medication during longitudinal follow-up, given that the authors were not directly involved in the clinical management of most 22q11DS participants. As such, we cannot draw confident conclusions on the potential influence of psychotropic medication prescribed after-baseline assessment on longitudinal BMI trajectories, especially considering that the effects of psychostimulant medication on BMI, are tightly related to treatment compliance(25, 26).

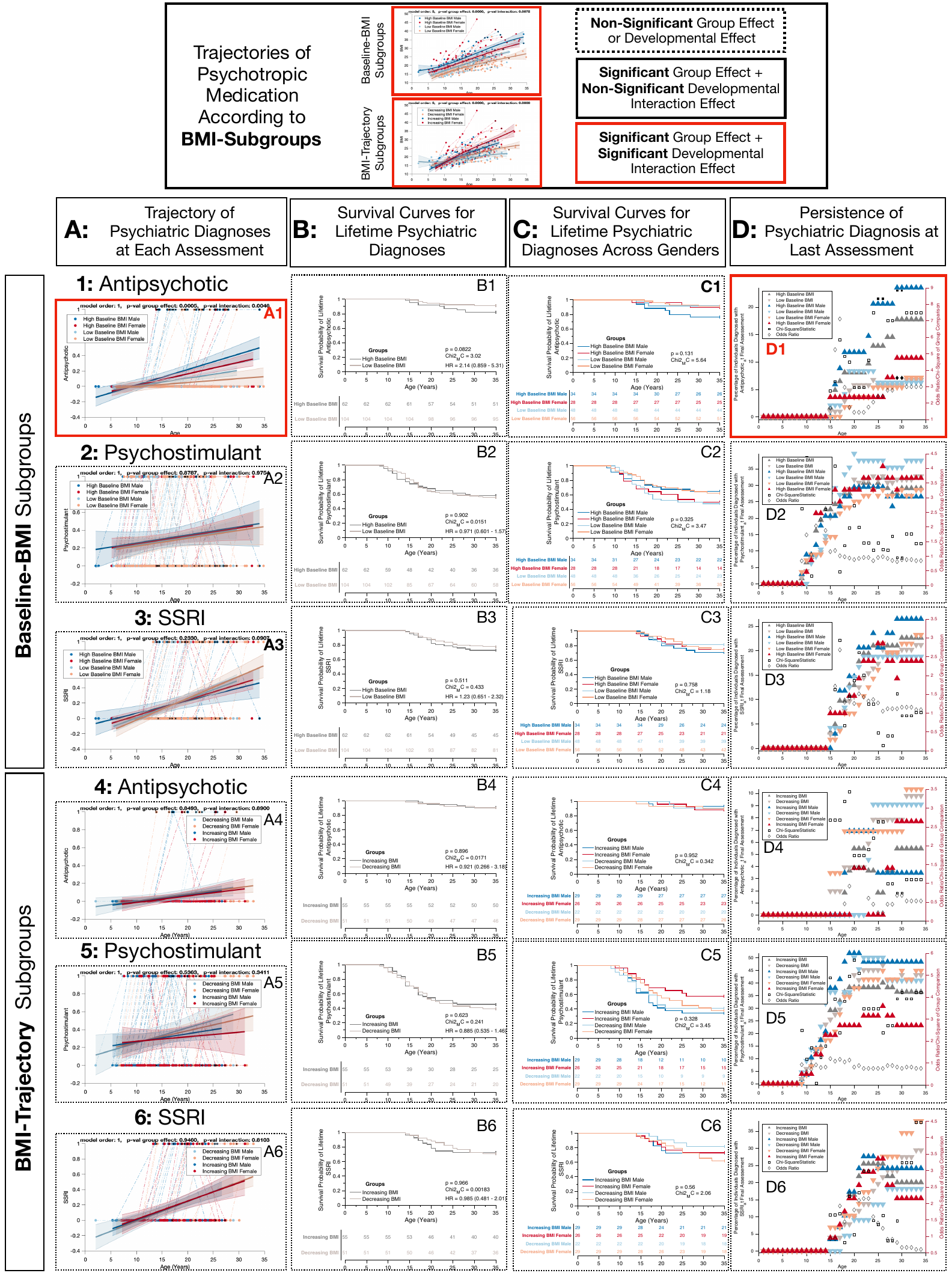

Supplementary Figure 2: Psychotropic Medication Use Across BMI Subgroups

Trajectories of use of psychotropic medication, compared across subgroups of 22q11DS individuals separated according to Baseline-BMI in Panels 1-3 High-Baseline-BMI-Males in dark blue, High-Baseline-BMI-Females in red, Low-Baseline-BMI-Males in light blue, and Low-Baseline-BMI-Females in orange. and according to BMI-Trajectory in Panels 4-6 Increasing BMI-Males in dark blue, Increasing-BMI-Females in red, Decreasing-BMI-Males in light blue, and Decreasing-BMI-Females in orange. Figures displaying significant group differences are outlined by solid black lines. Figures displaying significant developmental differences are outlined by solid red lines. Panels A1:A6: Trajectories of psychotropic medication use at individual assessments, compared across BMI subgroups using MMLR. **Panels B1:B6:** Kaplan-Meier survival curves for lifetime use of psychotropic medication**. Panels C1:C6:** Kaplan-Meier survival curves for lifetime use of psychotropic medication across gender subgroups. Panels D1:D6: Prevalence of use of psychotropic medication persisting until the last follow-up assessment.

##### 2.4 Comparison of DSM-5 Psychiatric Diagnoses across BMI Subgroups

Here we report comparisons of DSM-V diagnosis of psychotic disorders, mood disorders, anxiety disorders, and ADHD across BMI Subgroups. ADHD diagnoses that were separated in purely Inattentive-ADD and Combined-ADHD subtypes, according to the presence of significant Impulsivity/Hyperactivity traits, defined as >3 Impulsivity/Hyperactivity symptoms. The presence of a psychiatric diagnosis at each individual assessment was compared across subgroups using Mixed-Models Linear Regression. Results are reported in Panels A of Supplementary Figures 3 and 4. Moreover we constructed Kaplan-Meyer survival curves by measuring the lifetime prevalence of discrete outcomes across subgroups, considering assessments performed at increasing ages, in successive 1-year iterations which we compared across BMI-subgroups, using log rank test, implemented in MATLAB (<https://github.com/aebergl/MatSurv>). Results of Kaplan-Meyer survival curve analysis are reported in Panels B of Supplementary Figures 3 and 4 for merged gender subgroups and in Panels C considering genders separately. We also measured the prevalence of psychiatric diagnoses that persisted until the end of longitudinal follow-up, which we compared across BMI-subgroups using a 2-sample chi-square test, when ignoring the effect of gender, and a 4-sample chi-square test, when considering gender subgroups separately. Results are reported in Panels D of Supplementary Figures 3 and 4.

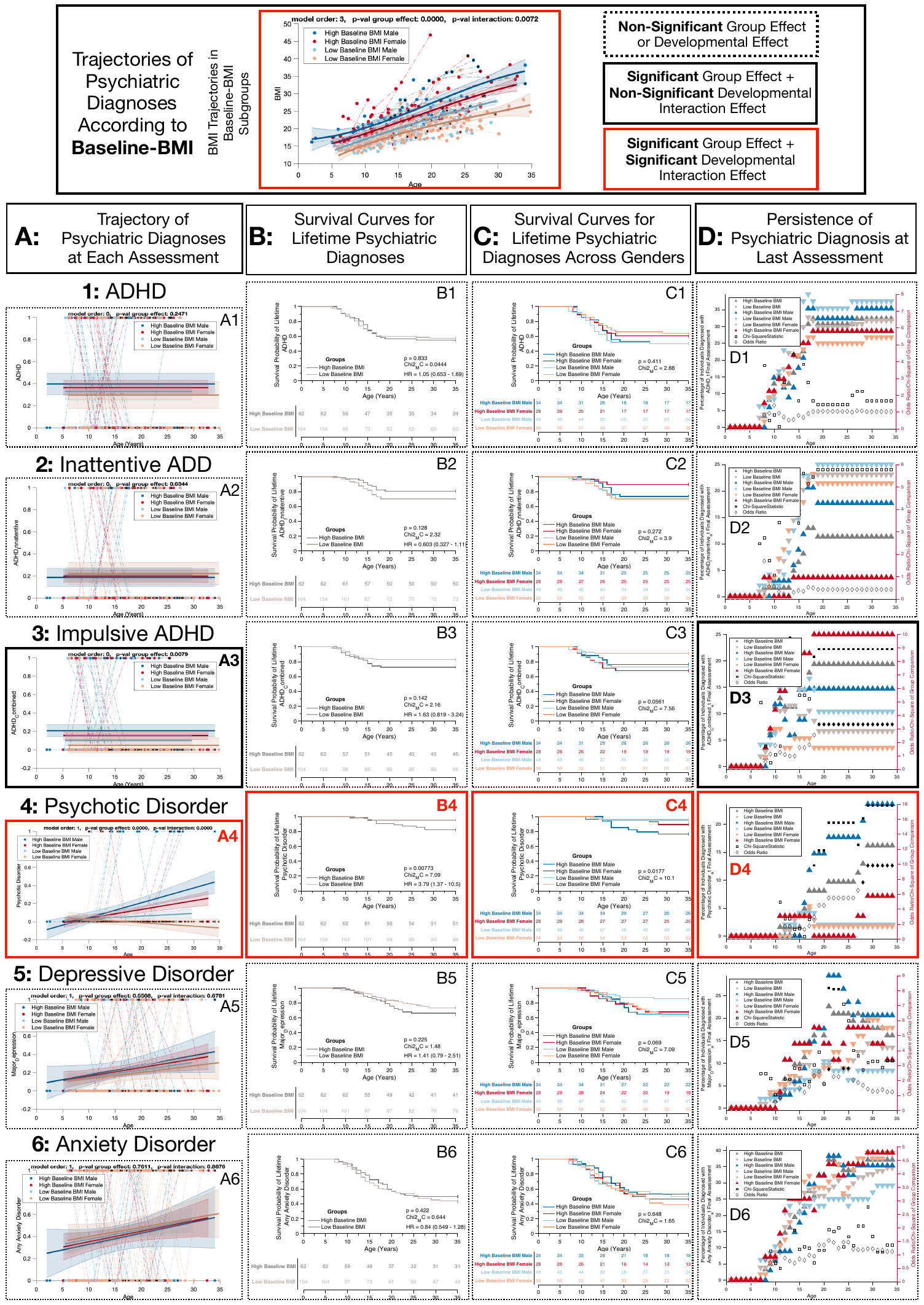

Supplementary Figure 3: Psychiatric Diagnoses across Baseline BMI Subgroups

Trajectories of main psychiatric diagnoses, compared across subgroups of 22q11DS individuals separated according to Baseline BMI. High-Baseline-BMI-Males in dark blue, High-Baseline-BMI-Females in red, Low-Baseline-BMI-Males in light blue, and Low-Baseline-BMI-Females in orange. Figures displaying significant group differences are outlined by solid black lines. Figures displaying significant developmental differences are outlined by solid red lines. **Panels A1:A6:** Trajectories of psychiatric diagnoses at individual assessments, compared across Baseline-BMI subgroups using MMLR. **Panels B1:B6:** Kaplan-Meier survival curves for lifetime psychiatric diagnoses. **Panels C1:C6:** Kaplan-Meier survival curves for lifetime psychiatric diagnoses across gender subgroups. **Panels D1:D6:** Prevalence of psychiatric diagnosis persisting until last follow-up assessment. **Panels A1:D1:** ADHD. **Panels A2:D2:** Purely Inattentive ADHD. **Panels A3:D3:** ADHD with combined Impulsivity. **Panels A4:D4:** Psychotic Disorders. **Panels A5:D5:** Depressive Disorders. **Panels A5:D5:** Anxiety Disorders

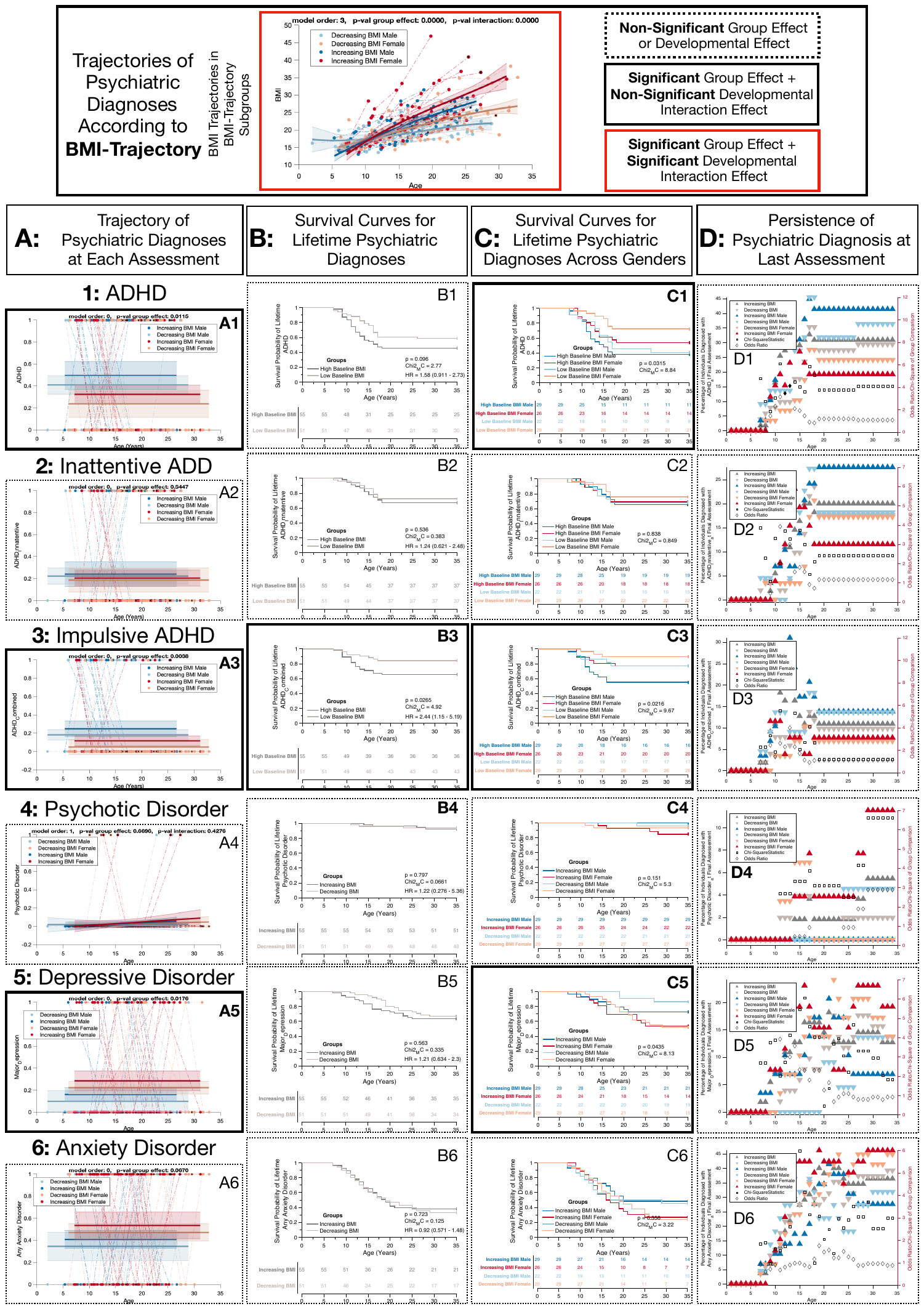

Supplementary Figure 4: : Psychiatric Diagnoses across BMI-Trajectory Subgroups

Trajectories of main psychiatric diagnoses, compared across subgroups of 22q11DS individuals separated according to BMI Trajectory. Increasing BMI-Males in dark blue, Increasing-BMI-Females in red, Decreasing-BMI-Males in light blue, and Decreasing-BMI-Females in orange. Figures displaying significant group differences are outlined by solid black lines. Figures displaying significant developmental differences are outlined by solid red lines. **Panels A1:A6:** Trajectories of psychiatric diagnoses at individual assessments, compared across Baseline-BMI subgroups using MMLR. **Panels B1:B6:** Kaplan-Meier survival curves for lifetime psychiatric diagnoses. **Panels C1:C6:** Kaplan-Meier survival curves for lifetime psychiatric diagnoses across gender subgroups. **Panels D1:D6:** Prevalence of psychiatric diagnosis persisting until last follow-up assessment. **Panels A1:D1:** ADHD. **Panels A2:D2:** Purely Inattentive ADHD. **Panels A3:D3:** ADHD with combined Impulsivity. **Panels A4:D4:** Psychotic Disorders. **Panels A5:D5:** Depressive Disorders. **Panels A5:D5:** Anxiety Disorders

##### 2.5 Comparison of Trajectories of individual Clinical/Neurocognitive Scores across BMI Subgroups

Here we report the clinical trajectories of individual clinical and neurocognitive scores compared across BMI trajectory and Baseline-BMI subgroups using Mixed-Model-Linear-Regression (MMLR). Additionally, across Baseline-BMI subgroups clinical trajectories are modelled according to time from baseline assessment, in order to investigate whether worsting of psychiatric difficulties associated to High-Baseline-BMI would be better predicted by a dose-effect relationship with duration of High-BMI status than by age. We employed False-Discovery-Rate to correct for multiple comparisons of different items in each psychometric instrument. Results of MMLR for each score and comparison are reported in Supplementary Table 2 and in Supplementary Figures 3 to 8.

| **Supplementary Table 1:**  **Clinical and Neurocognitive Trajectories Across BMI Subgroups** | | **Intercept/Age-Interaction** | **Increasing BMI** | **P-Value-Group-Effect/Interaction-Effect** | **Low Baseline BMI** | **High Baseline BMI** | **P-Value-Group-Effect/Interaction-Effect** | **P-Value-Group-Effect/ Interaction-Effect Time From Baseline Assessment** |
| --- | --- | --- | --- | --- | --- | --- | --- | --- |
| **BPRS Clinical Trajectories** | Total Score | 39.6/NaN | 40.5/NaN | 0.82/NaN | 40.3/NaN | 44.7/NaN | 0.0042/NaN | 0.0042/NaN |
|  | Anxiety | 2.58/NaN | 2.61/NaN | 0.243/NaN | 2.68/NaN | 2.87/NaN | 0.131/NaN | 0.131/NaN |
|  | Bizarre Behavior | 1.38/NaN | 1.3/NaN | 0.329/NaN | 1.12/0.0129 | 0.919/0.0417 | 0.0012/0.0208 | 0/0.0004 |
|  | Blunted Affect | 2.81/NaN | 2.72/NaN | 0.147/NaN | 2.74/NaN | 2.86/NaN | 0.323/NaN | 0.323/NaN |
|  | Conceptual Disorganization | 3.02/-0.0428 | 3.12/-0.045 | 0.907/0.873 | 3.07/-0.0428 | 2.73/-0.0108 | 0.0272/0.0487 | 0.0002/0.0001 |
|  | Depression | 1.45/NaN | 1.83/NaN | 0.0003/0.0058 | 1.65/NaN | 1.89/NaN | 0.0486/NaN | 0.0486/NaN |
|  | Disorientation | 1.05/0.0392 | 1.35/0.0211 | 0.473/0.221 | 1.3/0.0251 | 1.25/0.0416 | 0.044/0.227 | 0.0036/0.0143 |
|  | Distractibility | 3.5/-0.0286 | 3.9/-0.0537 | 0.278/0.111 | 3.57/-0.0335 | 3.8/-0.0379 | 0.249/0.743 | 0.352/0.948 |
|  | Elevated Mood | 1.1/NaN | 1.17/NaN | 0.005/0.0025 | 1.14/NaN | 1.32/NaN | 0.003/NaN | 0.003/NaN |
|  | Emotional Withdrawal | 2.63/NaN | 2.53/NaN | 0.669/NaN | 2.56/NaN | 2.72/NaN | 0.261/NaN | 0.261/NaN |
|  | Excitement | 1.37/-0.0102 | 1.78/-0.032 | 0.112/0.0449 | 1.21/NaN | 1.39/NaN | 0.0084/NaN | 0.0084/NaN |
|  | Grandiosity | 1.09/NaN | 1.08/NaN | 0.754/NaN | 1.07/NaN | 1.19/NaN | 0.012/NaN | 0.012/NaN |
|  | Guilt | 1.13/NaN | 1.39/NaN | 0.0013/0.0578 | 1.25/NaN | 1.5/NaN | 0.0037/NaN | 0.0037/NaN |
|  | Hallucinations | 1.62/NaN | 1.81/NaN | 0.347/NaN | 1.69/NaN | 2.05/NaN | 0.0081/NaN | 0.0081/NaN |
|  | Hostility | 1.04/NaN | 1.07/NaN | 0.354/NaN | 1.16/NaN | 1.18/NaN | 0.769/NaN | 0.769/NaN |
|  | Mannerisms | 1.16/NaN | 1.16/NaN | 0.183/NaN | 1.2/NaN | 1.3/NaN | 0.122/NaN | 0.122/NaN |
|  | Motor Hyperactivity | 1.85/-0.0326 | 2.07/-0.0456 | 0.479/0.227 | 1.89/-0.0333 | 1.99/-0.0327 | 0.278/0.95 | 0.36/0.476 |
|  | Motor Retardation | 1.45/NaN | 1.49/NaN | 0.39/NaN | 1.54/-0.00277 | 0.928/0.0358 | 0.0025/0.0007 | 0.0003/0.0001 |
|  | Self-Neglect | 0.923/0.046 | 0.846/0.0544 | 0.733/0.575 | 0.88/0.0486 | 0.937/0.0611 | 0.0244/0.356 | 0.0015/0.0162 |
|  | Somatic Problems | 1.14/0.0233 | 0.902/0.0334 | 0.568/0.469 | 0.892/0.0339 | 1.39/0.0208 | 0.0087/0.298 | 0.0069/0.782 |
|  | Suicidality | 1.08/NaN | 1.2/NaN | 0.0237/NaN | 1.15/NaN | 1.37/NaN | 0.0042/NaN | 0.0042/NaN |
|  | Suspiciousness | 1.84/NaN | 2.07/NaN | 0.0744/NaN | 1.46/0.0301 | 1.83/0.0243 | 0.0684/0.684 | 0.0252/NaN |
|  | Tension | 1.66/NaN | 1.57/NaN | 0.629/NaN | 1.65/NaN | 1.78/NaN | 0.132/NaN | 0.21/0.386 |
|  | Uncooperativeness | 1.22/NaN | 1.16/NaN | 0.614/NaN | 1.19/NaN | 1.33/NaN | 0.098/NaN | 0.098/NaN |
|  | Unsual Thought Content | 1.53/NaN | 1.65/NaN | 0.351/NaN | 1.6/NaN | 1.88/NaN | 0.0308/NaN | 0.006/0.0203 |
| **C/ABCL Clinical Trajectories**  **T-Scores** | Total Problems | 60.8/-0.107 | 69.1/-0.426 | 0.0043/0.0227 | 60.2/NaN | 64/NaN | 0.0028/NaN | 0.0028/NaN |
|  | Internalizing Problems | 61.8/NaN | 63.2/NaN | 0.372/NaN | 62/NaN | 65.5/NaN | 0.0072/NaN | 0.0072/NaN |
|  | Externalizing Problems | 51.9/NaN | 55.4/NaN | 0.0137/NaN | 53.5/NaN | 56.5/NaN | 0.0129/NaN | 0.0129/NaN |
|  | Thought Problems | 58.8/NaN | 60.3/NaN | 0.278/NaN | 60.3/NaN | 62.6/NaN | 0.0713/NaN | 0.0713/NaN |
|  | Anxiety/Depression Problems | 60/NaN | 62.6/NaN | 0.0587/NaN | 61/NaN | 64.5/NaN | 0.0053/NaN | 0.0053/NaN |
|  | Somatic Problems | 59.4/NaN | 61.5/NaN | 0.091/NaN | 60.5/NaN | 61.7/NaN | 0.286/NaN | 0.286/NaN |
|  | Social Problems | 67.9/-0.519 | 74.5/-0.777 | 0.0258/0.0801 | 68.9/-0.53 | 70.7/-0.475 | 0.0369/0.661 | 0.0349/0.511 |
|  | Withdrawn Problems | 62.7/NaN | 63.8/NaN | 0.445/NaN | 63.6/NaN | 64.9/NaN | 0.283/NaN | 0.283/NaN |
|  | Behavioural Agitation | 63/NaN | 65.5/NaN | 0.0034/NaN | 64.6/NaN | 65.5/NaN | 0.0089/NaN | 0.0089/NaN |
|  | Attention Problems | 54.1/NaN | 57/NaN | 0.0546/NaN | 55.7/NaN | 58.2/NaN | 0.391/NaN | 0.391/NaN |
| **SIPS Clinical Trajectories** | Unusual Thought Content | 1.11/-0.0196 | 1.1/0.0004 | 0.141/0.462 | 1.07/-0.00676 | 0.71/0.0337 | 0.0212/0.0931 | 0.004/0.0141 |
|  | Persecutory Ideas | 1.49/-0.0255 | 1.45/-0.00687 | 0.147/0.462 | 1.33/-0.00796 | 1.18/0.0228 | 0.0116/0.155 | 0.0135/0.174 |
|  | Grandiosity | -0.0593/0.00924 | -0.068/0.00708 | 0.709/0.816 | -0.208/0.0153 | -0.126/0.0173 | 0.0831/0.797 | 0.0131/0.0325 |
|  | Perceptual Abnormalities | 2.38/-0.0636 | 3.3/-0.0966 | 0.216/0.347 | 2.68/-0.0724 | 2.2/-0.0244 | 0.0291/0.0946 | 0.0105/0.0263 |
|  | Disorganized Communication | 0.464/-0.00149 | 0.32/0.00173 | 0.769/0.847 | 0.27/0.00472 | 0.513/0.0114 | 0.0099/0.684 | 0.0006/0.02 |
|  | Social Anhedonia | 1.78/0.0145 | 1.25/0.0455 | 0.521/0.255 | 1.6/0.0286 | 1.9/0.0127 | 0.776/0.477 | 0.867/0.594 |
|  | Avolition | 1.59/0.0285 | 0.956/0.0584 | 0.451/0.266 | 1.07/0.0544 | 1.31/0.056 | 0.181/0.939 | 0.0959/0.244 |
|  | Expression Emotion | -1.01/0.366 | 1.73/-0.00423 | 0.0244/0.0142 | 2.21/-0.0111 | 1.86/0.00695 | 0.679/0.382 | 0.423/0.191 |
|  | Experience Emotion | 0.151/0.0237 | 0.263/0.0163 | 0.935/0.731 | 0.222/0.0164 | 0.283/0.0283 | 0.0653/0.511 | 0.0503/0.392 |
|  | Ideational Richness | 4.42/-0.0486 | 3.96/-0.0297 | 0.618/0.483 | 4.34/-0.0494 | 4.06/-0.0252 | 0.3/0.257 | 0.0463/0.025 |
|  | Occupational Functioning | 2.57/-0.0208 | 1.5/0.0433 | 0.0313/0.0088 | 1.86/0.0195 | 2.18/0.0199 | 0.0826/0.983 | 0.0393/0.168 |
|  | Odd Behaviour | -0.0153/0.0337 | -0.00425/0.0262 | 0.596/0.667 | 0.16/0.0181 | 0.000953/0.0472 | 0.0026/0.0835 | 0.0001/0.0017 |
|  | Bizarre Thinking | -0.158/0.0247 | 0.127/0.00886 | 0.603/0.316 | 0.257/0.00364 | -0.474/0.0578 | 0.0001/0.0004 | 0/0 |
|  | Attention Diffuculties | 2.68/-0.00151 | 2.41/0.00993 | 0.796/0.596 | 2.55/0.00537 | 2.72/0.00725 | 0.288/0.913 | 0.186/0.397 |
|  | Impaired Personal Hygiene | 0.576/0.0185 | 0.815/0.00923 | 0.782/0.648 | 0.695/0.00981 | 0.541/0.0342 | 0.0181/0.144 | 0.0031/0.0219 |
|  | Sleep Disturbances | 0.713/0.024 | 1.15/0.0097 | 0.543/0.613 | 0.953/0.016 | 0.598/0.047 | 0.158/0.173 | 0.268/0.447 |
|  | Dysphoric Mood | 0.582/0.00332 | 0.107/0.0585 | 0.0016/0.0387 | 0.35/0.0389 | 0.676/0.0276 | 0.712/0.635 | 0.516/0.326 |
|  | Motor Disturbances | 0.0333/0.0165 | 0.179/0.017 | 0.448/0.983 | 0.403/-0.000505 | -0.224/0.0441 | 0.0016/0.0028 | 0.0003/0.0004 |
|  | Impaired Tollerance to Stress | -0.104/0.0742 | 0.317/0.0465 | 0.491/0.267 | 0.21/0.0565 | 0.111/0.0671 | 0.708/0.625 | 0.0555/0.0249 |
| **IQ and CPT Neurocognitive Trajecotories** | Full-Scale IQ | 71.6/NaN | 72/NaN | 0.851/NaN | 72.1/NaN | 69.3/NaN | 0.121/NaN | 0.121/NaN |
|  | Verbal IQ | 85/-0.468 | 83/-0.374 | 0.825/0.553 | 82.9/-0.328 | 85.1/-0.531 | 0.305/0.176 | 0.242/0.154 |
|  | Performance IQ | 72.3/NaN | 73.3/NaN | 0.612/NaN | 73/NaN | 69.6/NaN | 0.0478/NaN | 0.0478/NaN |
|  | Processing Speed IQ | 81.2/NaN | 81.9/NaN | 0.836/NaN | 82.5/NaN | 76.4/NaN | 0.0123/NaN | 0.0123/NaN |
|  | CPT Omission Errors T-Score | 63.1/-1.36 | 40.5/1.46 | 0.184/0.285 | 48.8/0.483 | 30.4/2.1 | 0/0.0001 | 0/0.0002 |
|  | CPT Commission Errors T-Score | 26.2/3.13 | 40.5/1.35 | 0.228/0.327 | 43.8/0.722 | 52.5/0.056 | 0.0148/0.0101 | 0.396/0.332 |
|  | CPT Commission/Omission Errors | 39.6/7.68 | 101/0.0707 | 0.0139/0.0546 | 93.8/0.495 | 123/-1.96 | 0.0001/0.0006 | 0.0001/0.0002 |
|  | CPT Hit Reaction Time T-Scores | 95.2/-5.71 | 60.8/-1.71 | 0.0111/0.0161 | 58.2/-0.867 | 48.9/-0.0199 | 0.0006/0.0044 | 0.0076/0.0192 |

Supplementary Table 1: Differences in clinical trajectories of clinical and neurocognitive scores across BMI Subgroups

Differences in clinical trajectories of clinical and neurocognitive scores tested with Mixed Model Linear Regression (MMLR) across Increasing vs Decreasing BMI Trajectory Subgroups and High vs Low Baseline BMI Subgroups. For each subgroup we report the sub-group specific beta values for intercept and age effect derived from the MMLR. For variables in which MMLR did not detect a significant overall age effect, group* age effect was not tested, and the corresponding beta and p values are reported as NaNs. Visual representation of the clinical trajectories of each variable are reported in Figure 2 and in supplementary material.

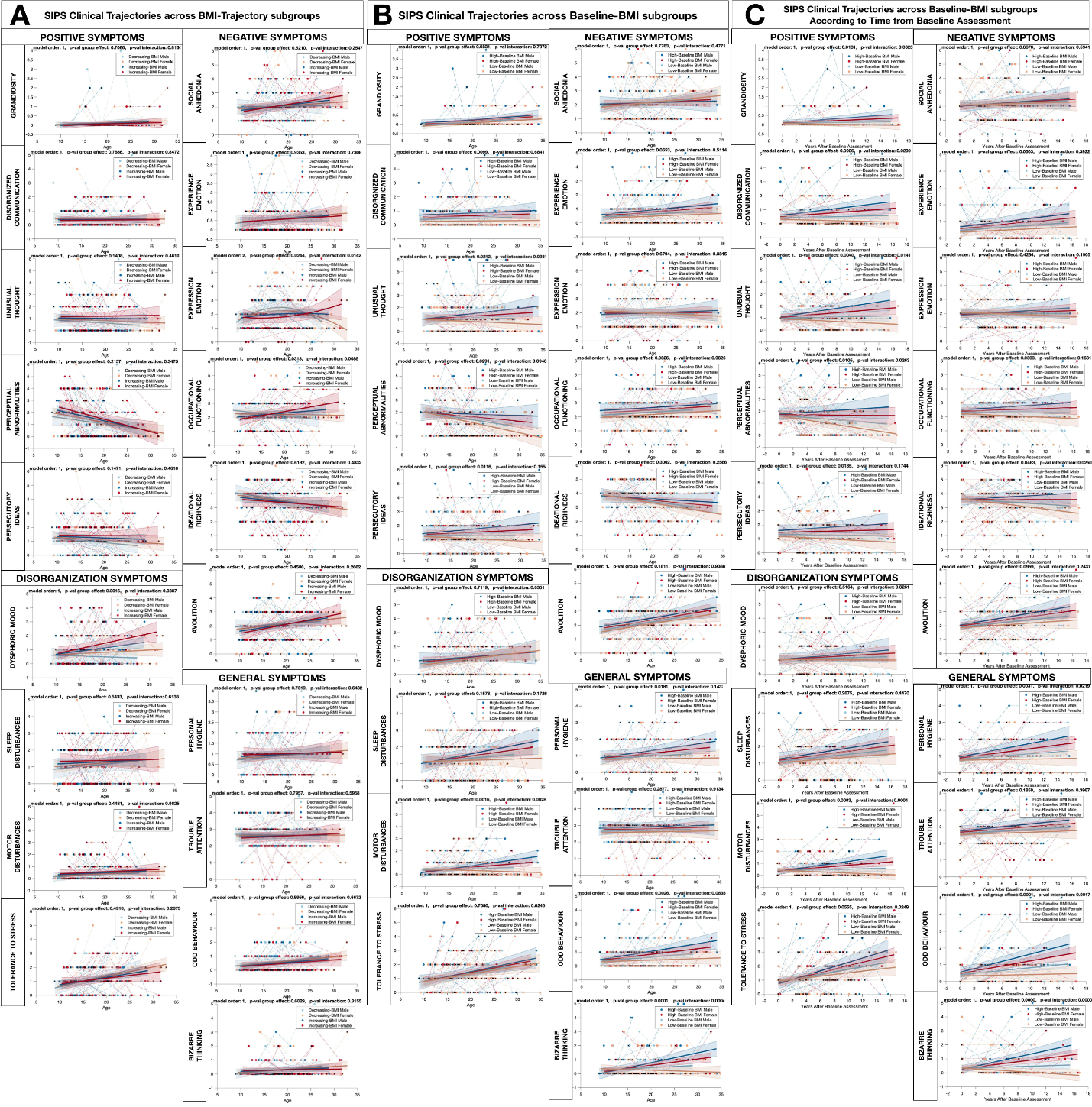

Supplementary Figure 5: Clinical Trajectories of items of the Structured Interview for Prodromal Syndromes

**Panel A**: Across BMI Trajectory Subgroups modelling for the effect of gender and yielding 4 subgroups: Increasing-BMI-Males in dark blue, Increasing-BMI-Females in red, Decreasing-BMI-Males in light blue, and Decreasing-BMI-Females in orange. **Panel B**: Across Baseline-BMI subgroups modelling for the effect of gender and yielding 4 subgroups: High-Baseline BMI-Males in dark blue, High-Baseline-BMI-Females in red, Low-Baseline-BMI-Males in light blue, and Low-Baseline-BMI-Females in orange. **Panel C**: Across Baseline-BMI subgroups according to Time from Baseline Assessment modelling for the effect of gender and yielding 4 subgroups: High-Baseline BMI-Males in dark blue, High-Baseline-BMI-Females in red, Low-Baseline-BMI-Males in light blue, and Low-Baseline-BMI-Females in orange.

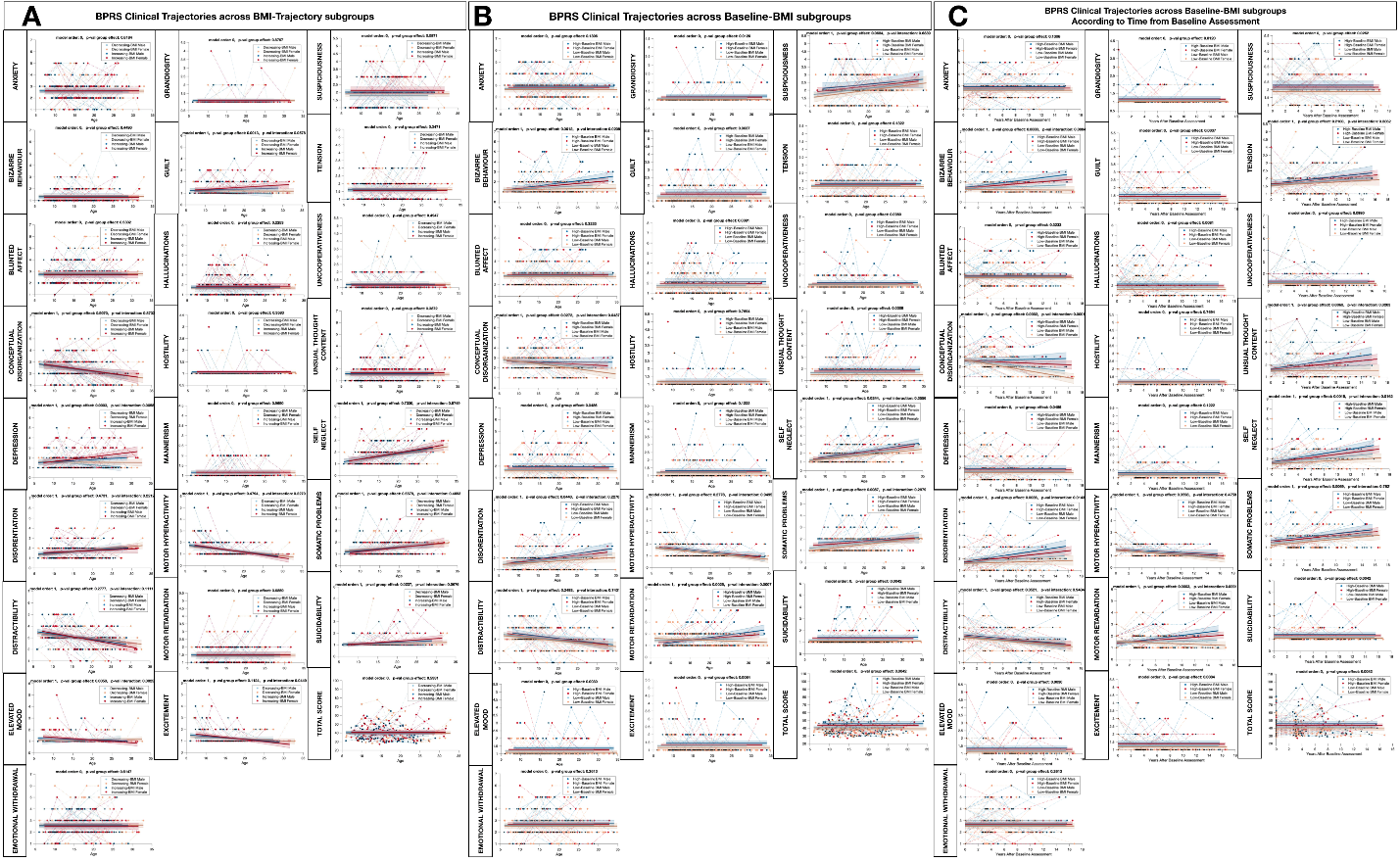

Supplementary Figure 6: Clinical Trajectories of items of the Brief Psychiatric Rating Scale

**Panel A**: Across BMI Trajectory Subgroups modelling for the effect of gender and yielding 4 subgroups: Increasing-BMI-Males in dark blue, Increasing-BMI-Females in red, Decreasing-BMI-Males in light blue, and Decreasing-BMI-Females in orange. **Panel B**: Across Baseline-BMI subgroups modelling for the effect of gender and yielding 4 subgroups: High-Baseline BMI-Males in dark blue, High-Baseline-BMI-Females in red, Low-Baseline-BMI-Males in light blue, and Low-Baseline-BMI-Females in orange. **Panel C**: Across Baseline-BMI subgroups according to Time from Baseline Assessment modelling for the effect of gender and yielding 4 subgroups: High-Baseline BMI-Males in dark blue, High-Baseline-BMI-Females in red, Low-Baseline-BMI-Males in light blue, and Low-Baseline-BMI-Females in orange.

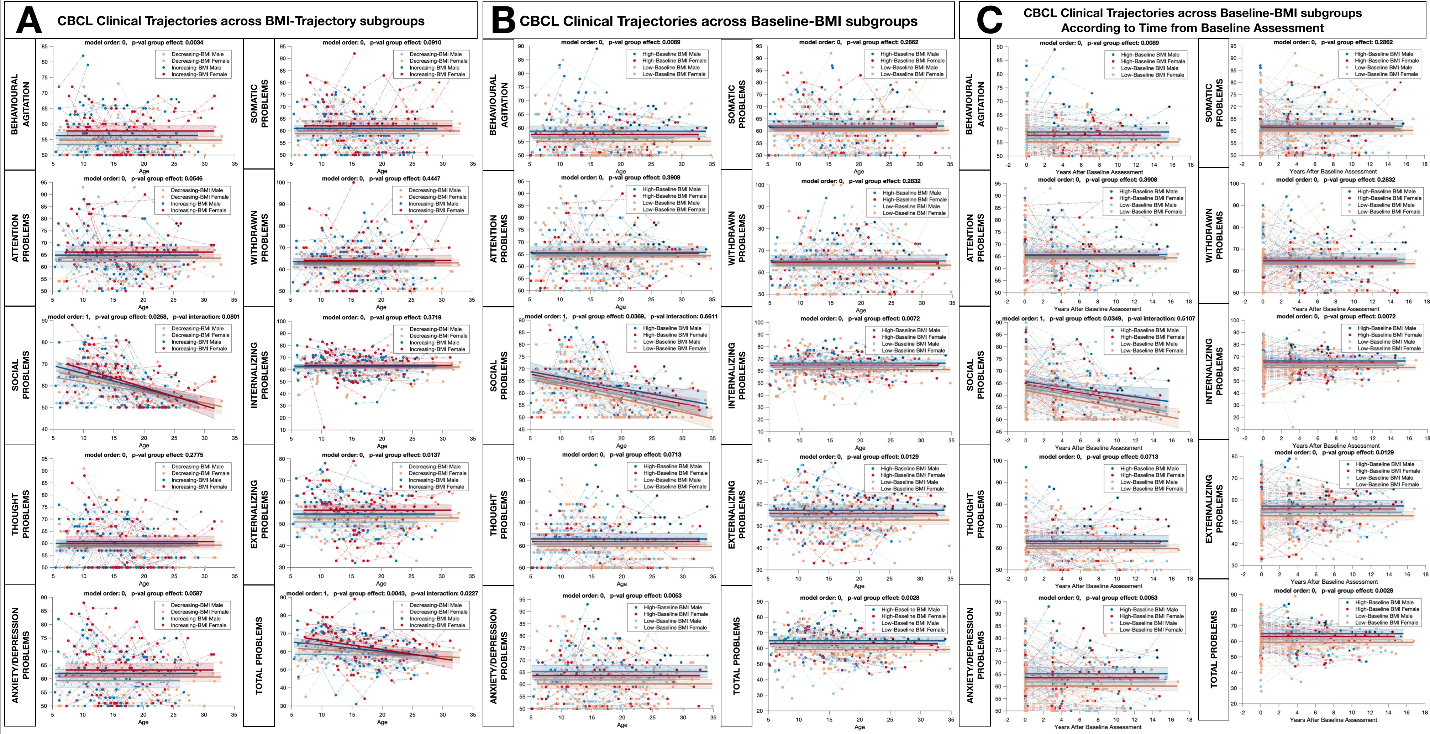

Supplementary Figure 7: Clinical Trajectories of items of the Child/Adult Behavioral Checklist

**Panel A**: Across BMI Trajectory Subgroups modelling for the effect of gender and yielding 4 subgroups: Increasing-BMI-Males in dark blue, Increasing-BMI-Females in red, Decreasing-BMI-Males in light blue, and Decreasing-BMI-Females in orange. **Panel B**: Across Baseline-BMI subgroups modelling for the effect of gender and yielding 4 subgroups: High-Baseline BMI-Males in dark blue, High-Baseline-BMI-Females in red, Low-Baseline-BMI-Males in light blue, and Low-Baseline-BMI-Females in orange. **Panel C**: Across Baseline-BMI subgroups according to Time from Baseline Assessment modelling for the effect of gender and yielding 4 subgroups: High-Baseline BMI-Males in dark blue, High-Baseline-BMI-Females in red, Low-Baseline-BMI-Males in light blue, and Low-Baseline-BMI-Females in orange.

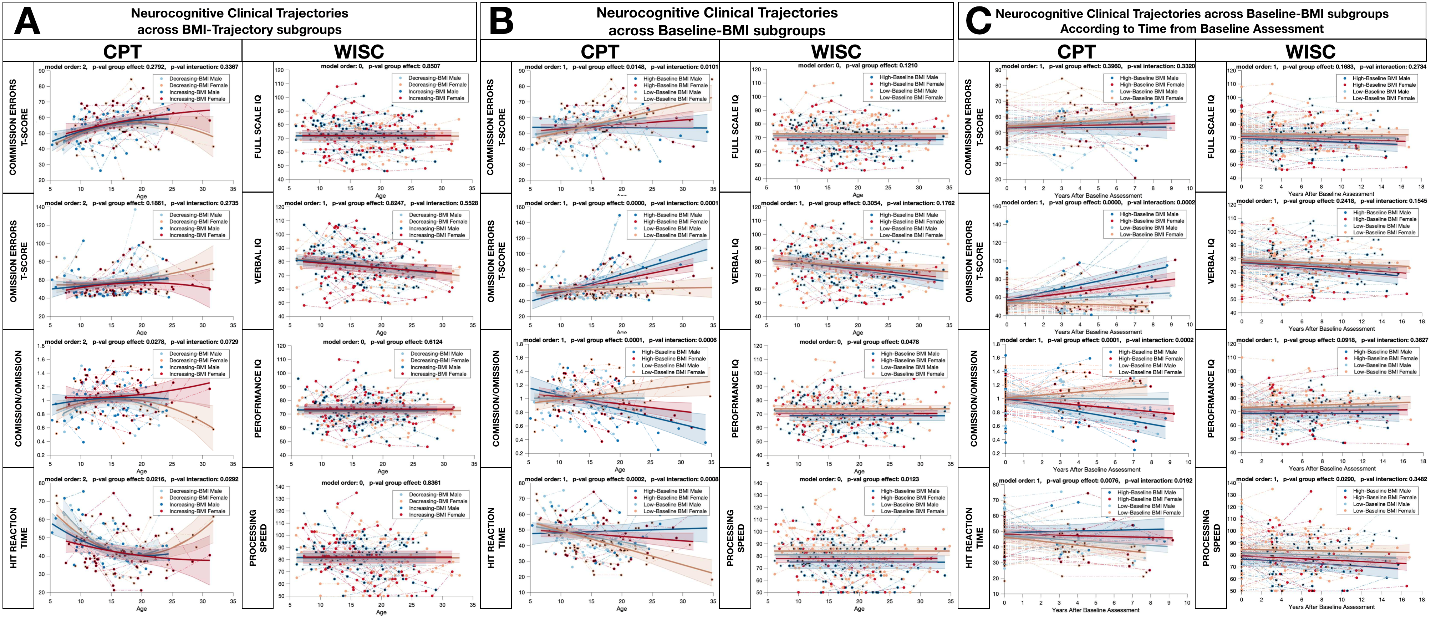

Supplementary Figure 8: Neurocognitive Trajectories

Neurocognitive Trajectories measured with the Child and Adult Version of Weschler Intelligence Scale and Connors Continuous Performance Test **Panel A**: Across BMI Trajectory Subgroups modelling for the effect of gender and yielding 4 subgroups: Increasing-BMI-Males in dark blue, Increasing-BMI-Females in red, Decreasing-BMI-Males in light blue, and Decreasing-BMI-Females in orange. **Panel B**: Across Baseline-BMI subgroups modelling for the effect of gender and yielding 4 subgroups: High-Baseline BMI-Males in dark blue, High-Baseline-BMI-Females in red, Low-Baseline-BMI-Males in light blue, and Low-Baseline-BMI-Females in orange. **Panel C**: Across Baseline-BMI subgroups according to Time from Baseline Assessment modelling for the effect of gender and yielding 4 subgroups: High-Baseline BMI-Males in dark blue, High-Baseline-BMI-Females in red, Low-Baseline-BMI-Males in light blue, and Low-Baseline-BMI-Females in orange.

##### 2.6 Comparison of Trajectories of Gray Matter Volume (GMV) across BMI Subgroups

Here we report the neurodevelopmental trajectories of Gray Matter Volume (GMV) in individual cerebellar and cortical subfields, compared across BMI trajectory and Baseline-BMI subgroups using Mixed-Model-Linear-Regression (MMLR). Additionally, across Baseline-BMI subgroups clinical trajectories are modelled according to time from baseline assessment, in order to investigate GMV reductions associated to High-Baseline-BMI would be better predicted by a dose-effect relationship with duration of High-BMI status than by age. Results of MMLR for each cerebellar and cortical subfield are reported in Supplementary Figure 9 and Supplementary Figure 10.

Supplementary Figure 9: Trajectories of Cerebellar Gray Matter Volume (GMV) across BMI Subgroups

Cerebellar trajectories compared across BMI subgroups for Total Cerebellum Gray Matter Volume and in 12 Bilateral Cerebellum Subfields.

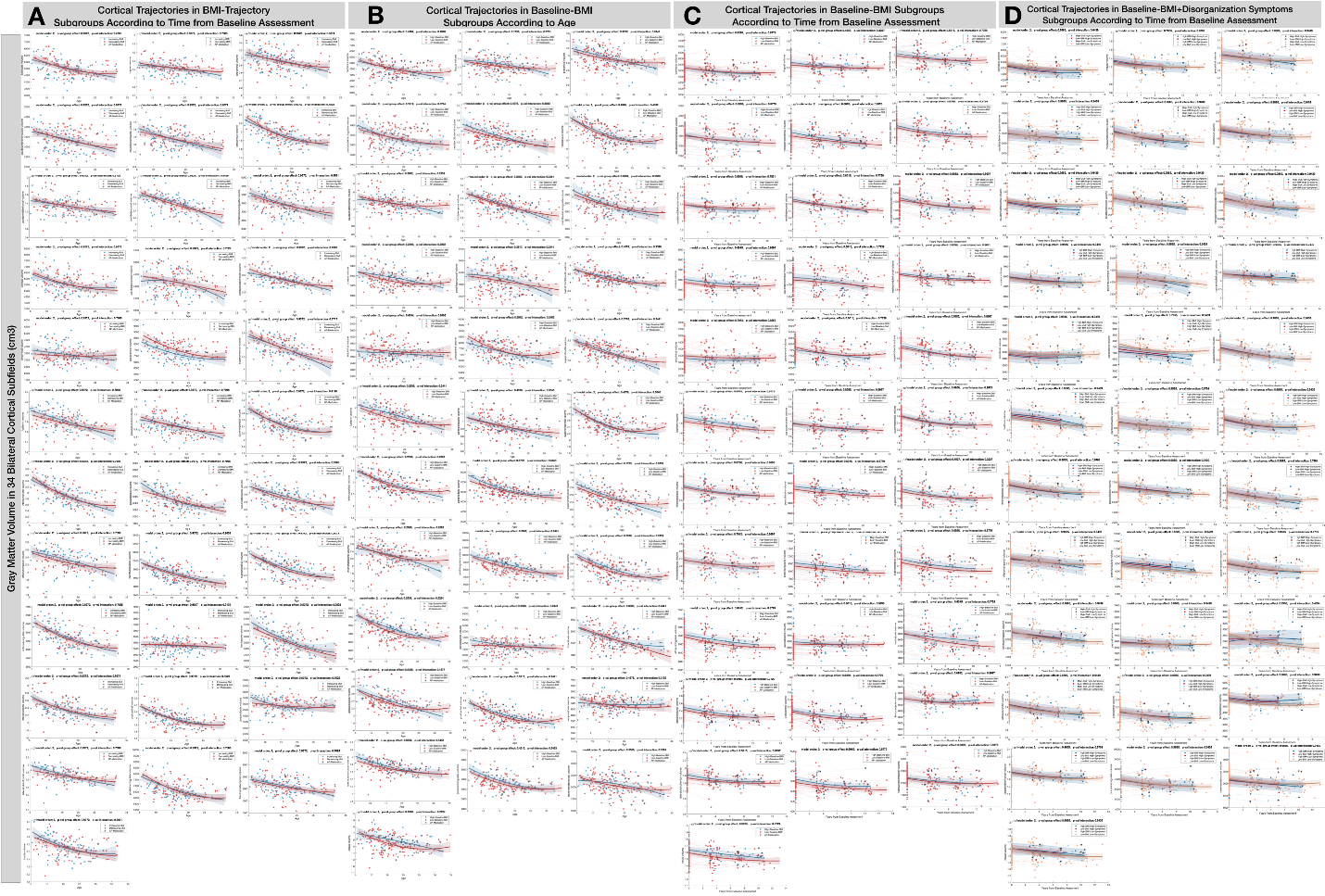

Supplementary Figure 10: Trajectories of Cortical Gray Matter Volume (GMV) across BMI Subgroups

Cortical trajectories compared across BMI subgroups for 34 Bilateral Cerebellum Subfields.

##### 2.7 PLCS Analysis of 3-way Clinical-Cerebellar-GMV-BMI Association

PLCS analysis revealed a component that accounts for the effect of SIPS-intensity diagnosis on the developmental trajectory of Cerebellum GMV. (P=0.002, R=0.27, See Supplementary Figure 11 Panels B-C). On the behavioral side, PLCS detected a negative effect of age on overall Cerebellum GMV, primarily driven by posterior-inferior cerebellum lobules. See Supplementary Figure 11 Panel A. Most SIPS-Diagnosis-by-Age variables loaded positively on this pattern, indicating that the age-related decline in posterior-inferior Cerebellum GMV was steeper in individuals with more severe symptom intensity. See Supplementary Figure 11 Panel D. To provide an alternative representation of this pattern, we derived a single multivariate Cerebellum-GVM score by performing matrix multiplication of the PLSC loading of cerebellum variable with GMV values measured at each longitudinal assessment. We then employed a mixed-model linear regression to identify which individual SIPS variables significantly moderate the trajectory of this Cerebellum-GVM Score, modelled both as a function of age and of Time-from-Baseline-Assessment (TBA). See Supplementary Figure 11. Results confirmed that individuals with at least moderate symptom intensity (>=3 on SIPS Scale), Cerebellum-GVM score showed sharper age-related decline with differences being even more striking when modelling Cerebellum-GVM trajectories as a function of TBA. SIPS variables that most significantly differentiated Cerebellum GMV trajectory were Bizarre Thinking, Odd Behavior, Disorganized Communication, Impaired Personal Hygiene, and Unusual Thought Content. See Supplementary Figure 12.

We then investigated whether Baseline-BMI status contributed to such Clinical-Cerebellar association. From the Clinical-Cerebellar PLCS analysis, we derived a **multivariate SIPS-Clinical Score** and a **Cerebellum-GMV Score** by matrix-multiplying the loadings of SIPS and Cerebellum-GMV variables with values measured at each longitudinal visit. Next, we tested whether BMI status differentially modulated the developmental trajectories of these multivariate scores. Results showed that **High-Baseline-BMI** contributed to both a sharper age-related increase in the **SIPS-Clinical Score** (P-Group=0.0026, P-Age-interaction=0.02, See Supplementary Figure 11 Panel E) and sharper age-related decline in the **Cerebellum-GMV Score** (P-Group=0.042, P-Age-interaction=0.034, See Supplementary Figure 11 Panel G). The divergence in both the **SIPS-Clinical Score** (P-Group-Effect<0.0001, P-Time-from-Baseline-interaction<0.0001) and **Cerebellum-GMV Score** (P-Group-Effect=0.016, P-Time-from-Baseline-interaction=0.01) was even more pronounced when modeling the effect of time from the baseline assessment. See Supplementary Figure 11 Panels E and H. This would suggest that **High-BMI status** may mediate the association between increasing SIPS intensity and the decline in **posterior-inferior Cerebellum-GMV**, through a dose-effect relationship with High-BMI status duration.

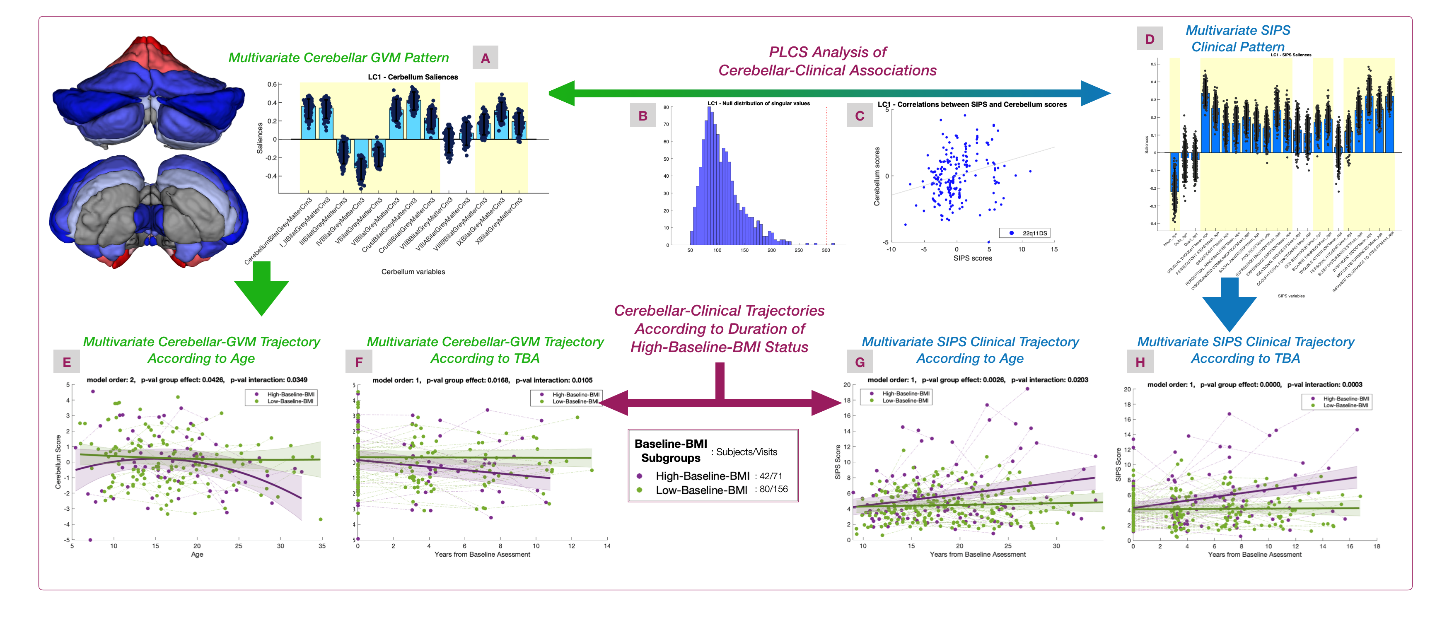

Supplementary Figure 11: PLCS Analysis of 3-way Clinical-Cerebellar-GMV-BMI Association

**Panel A:** Multivariate Cerebellar-GVM pattern reflecting Cerebellar-Clinical associations. Color coding in anatomical map reflects loading of individual cerebellar lobules, with blue indicating positive loading and red indicating negative loading. Total Cerebellar-GVM loaded positively on this pattern, driven by Posterior-Inferior Cerebellar lobules, while Anterior-Inferior lobules contributed mild negative loading. **Panel B:** Null-distribution derived from 1000 random permutations. **Panel C:** Association of Multivariate Cerebellar-GVM patterns and SIPS-Clinical patterns detected from PLCS analysis. **Panel D:** Multivariate SIPS clinical pattern, derived from PLCS analysis, capturing the impact of SIPS-Variables on age-related Cerebellar-GVM trajectories. The pattern detected a negative effect of age on Cerebellar-GVM. Most SIPS-Diagnosis-by-Age variables loaded positively onto this pattern, indicating that age-related Cerebellum-GMV reductions were steeper in individuals with higher symptom intensity. **Panel E:** Age-related trajectories of multivariate Cerebellar-GVM score derived from Cerebellar-Clinical PLCS analysis, according to High-vs-Low BMI status. **Panel F:** Trajectories of Multivariate Cerebellar-GVM score derived from Cerebellar-Clinical PLCS analysis, according to Duration of High-vs-Low BMI status. **Panel G:** Age-related trajectories of Multivariate SIPS-Clinical score derived from Cerebellar-Clinical PLCS analysis, according to Duration of High-vs-Low BMI status. **Panel H:** Trajectories of Multivariate SIPS-Clinical score derived from Cerebellar-Clinical PLCS analysis, according to duration of High-vs-Low BMI status.

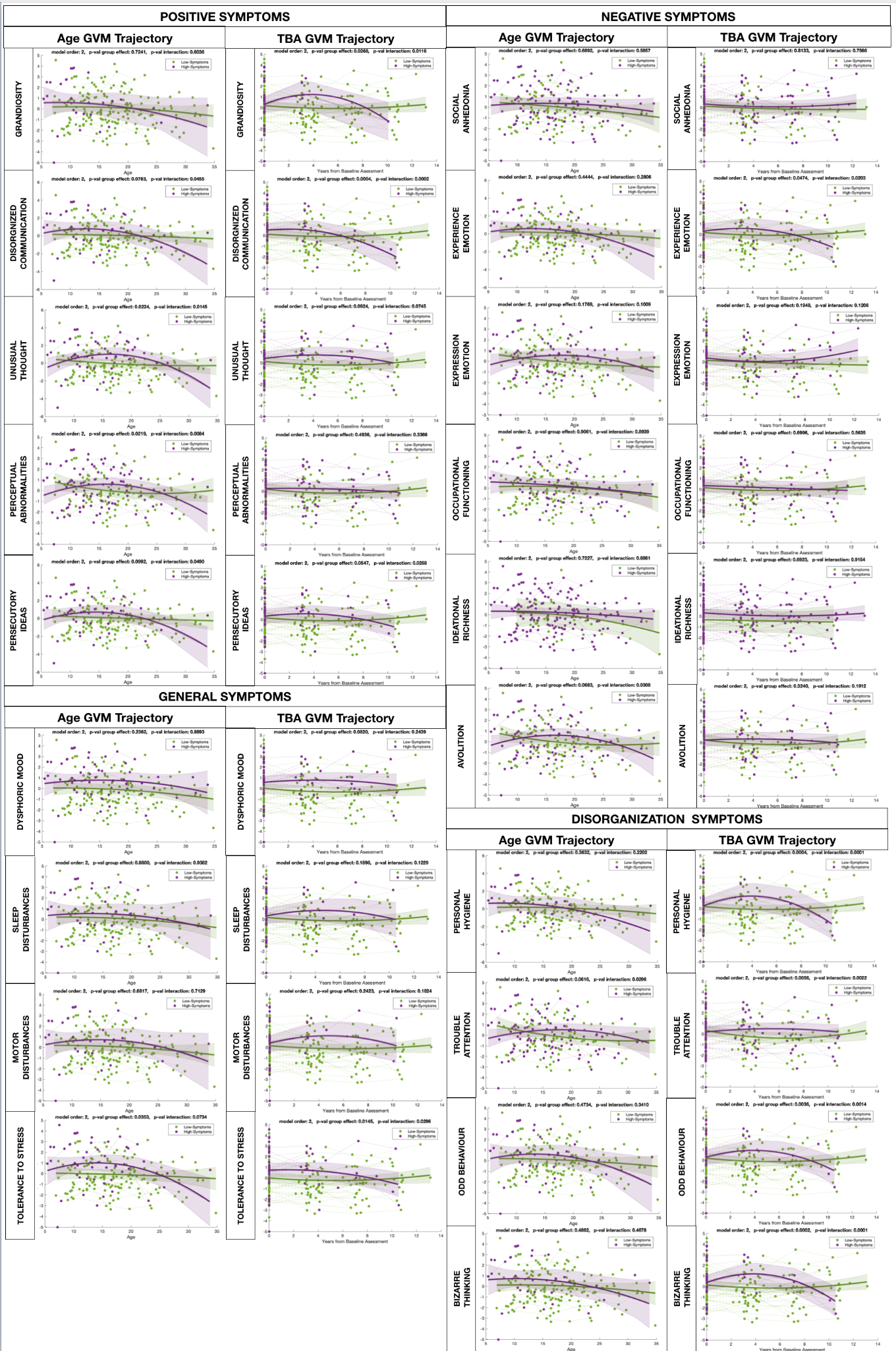

Supplementary Figure 12: Contribution of individual SIPS variables to Cerebellar-GMV trajectories captured by Clinical-Cerebellar PLSC analysis

A single multivariate Cerebellum-GVM score was derived through matrix multiplication of the PLSC loading of cerebellum variable with GMV values measured at each longitudinal assessment. We compared the trajectory of such Cerebellum-GVM scores as a function of both age and Time-from-Baseline-Assessment (TBA) across individuals divided according to the intensity (>3 on SIPS Scale) of individual SIPS symptoms. Individual symptoms that were significantly associated with atypical Cerebellum-GVM trajectory detected by PLCS are highlighted in purple.

##### 2.8 PLCS Analysis of 3-way Clinical-Cortical-GMV-BMI Association

To investigate the specificity of this findings to Cerebellum-GVM we the same PLCS analysis linking SIPS-intensity diagnosis with Cortical GMV which yielded 3 significant components. See Supplementary Figure 13. To provide a representation of the contribution of individual SIPS variables to this pattern we derived a Cortical-GVM score by performing matrix multiplication of the PLSC loading of cortical variables with GMV values measured at each longitudinal assessment. We compared the trajectory of each of the 3 Cortical GVM scores as a function across individuals divided according to the intensity (>3 on SIPS Scale) of individual symptoms. See Supplementary Figure 14.

The first Latent Component (LC1) primarily identified a negative effect of age on most cortical GVM values, along with significant loadings for most age-by-SIPS-diagnosis interactions. Unusual Thought Content showed the strongest individual contribution to this pattern, with more prolonged age-related GVM reductions persisting into adulthood. In contrast, these reductions tended to stabilize in individuals with fewer symptoms.

The second Latent Component (LC2) was characterized by positive loadings in the superior temporal, inferior frontal, parahippocampal, and posterior parietal regions, while sensory-motor and inferior temporal regions contributed negatively (highlighted in red; see Supplementary Figure 13). Negative symptoms, particularly Social Anhedonia and Avolition, contributed most strongly to this pattern. These symptoms were associated with early reductions in cortical GVM in positively loaded regions, with relatively little age-related reduction compared to individuals with less severe social anhedonia (see Supplementary Figure 14). General symptom variables such as Impaired Tolerance to Stress and Dysphoric Mood exerted an overall opposing influence on this pattern. However, when considered in isolation, no individual SIPS-diagnosis variable showed a significant association with the LC2 developmental pattern.

The third Latent Component (LC3) exhibited positive loadings in the superior frontal, superior temporal, and parahippocampal regions, while the middle and inferior temporal regions showed slight negative loadings (see Supplementary Figure 13). More severe general symptoms, particularly Sleep Disturbances and Impaired Tolerance to Stress, were associated with more pronounced age-related GVM reductions in positively loaded regions (see Supplementary Figure 14).

We then investigated whether Baseline-BMI status contributed to such Clinical-Cortical associations by testing whether BMI status differentially modulated the developmental trajectories of these multivariate clinical and Cortical-GVM scores derived from significant PLSC components. Interestingly High-BMI Status differentially moderated the clinical trajectory of SIPS-Behavioral scores derived from all 3 components (See Supplementary Figure 13 Panels G-H) but did not significantly affect developmental trajectory of Cortical-GVM scores derived from such patterns (See Supplementary Figure 13 Panels E-F). As such High-BMI status did not significantly mediate the association between clinical and Cortical-GVM-trajectories, which would suggest that the clinical correlates of High-BMI-status might be specifically related its effects on posterior-inferior Cerebellum GVM reductions.

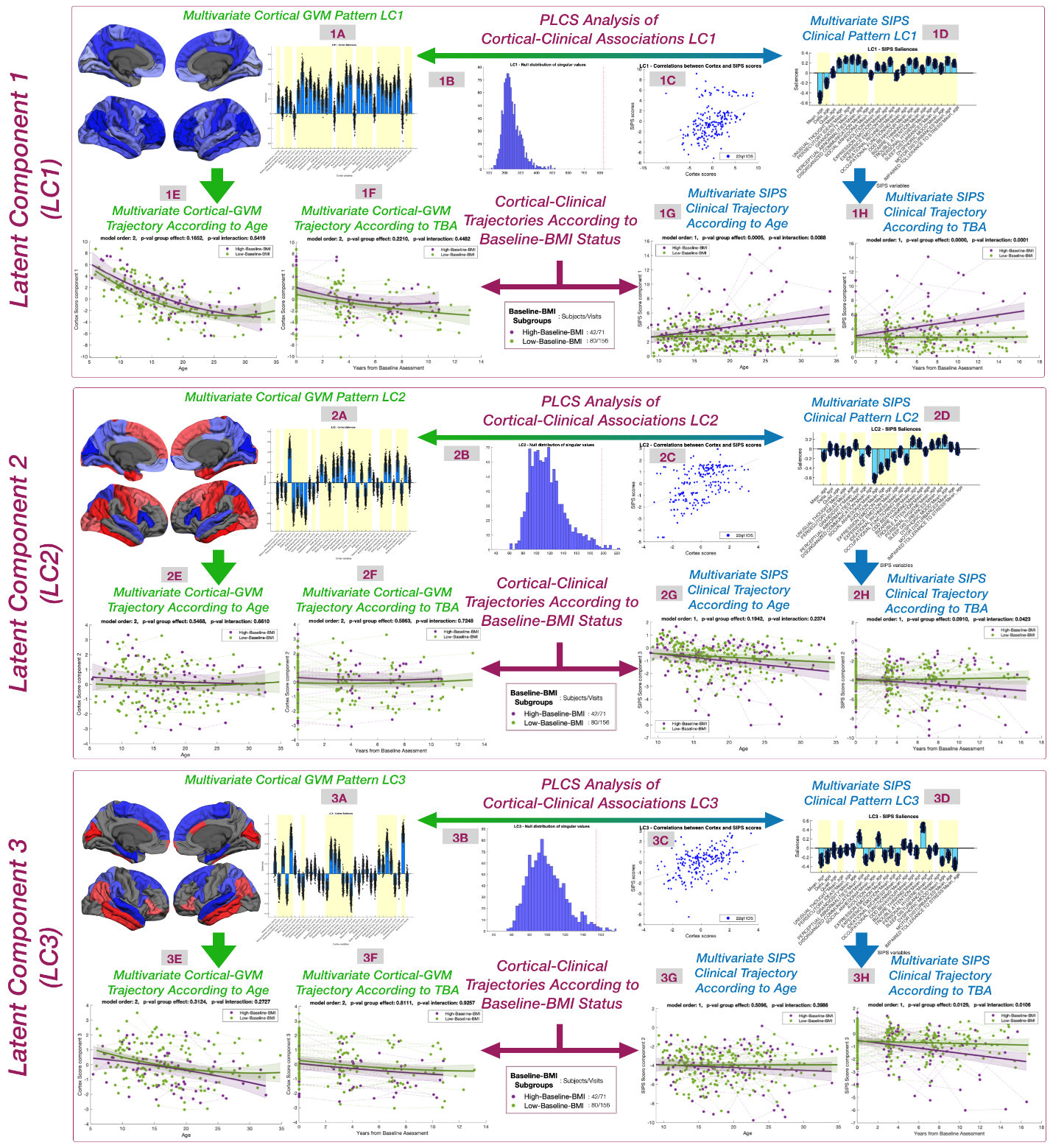

Supplementary Figure 13: PLSC Analysis of 3-way Clinical-Cortical-GMV-BMI Association

**Panels 1:** Results of first PLSC Latent Component (LC1). **Panels 2:** Results of second PLSC Latent Component (LC2). **Panels 3:** Results of third PLSC Latent Component (LC3). **Panels A-1-3:** Multivariate Cortical-GVM pattern reflecting Cortical-Clinical associations. Color coding in anatomical map reflects loading of individual cortical regions with blue indicating positive loading and red indicating negative loading. **Panels B-1-3:** Null-distribution derived from 1000 random permutations. **Panels C-1-3:** Association of Multivariate Cortical-GVM patterns and SIPS-Clinical patterns detected from PLCS analysis. **Panels D-1-3:** Multivariate SIPS clinical pattern, derived from PLCS analysis, capturing the impact of SIPS-Variables on age-related Cortical-GVM trajectories. **Panels E-1-3:** Age-related trajectories of multivariate Cortical-GVM score derived from Cortical-Clinical PLCS analysis, according to High-vs-Low BMI status. **Panels F-1-3:** **Trajectories** of Multivariate Cortical-GVM score derived from Cortical-Clinical PLCS analysis, according to Duration of High-vs-Low BMI status. **Panels G-1-3:** Age-related trajectories of Multivariate SIPS-Clinical score derived from Cortical-Clinical PLCS analysis, according to Duration of High-vs-Low BMI status. **Panels H-1-3:** Trajectories of Multivariate SIPS-Clinical score derived from Cortical-Clinical PLCS analysis, according to Duration of High-vs-Low BMI status.

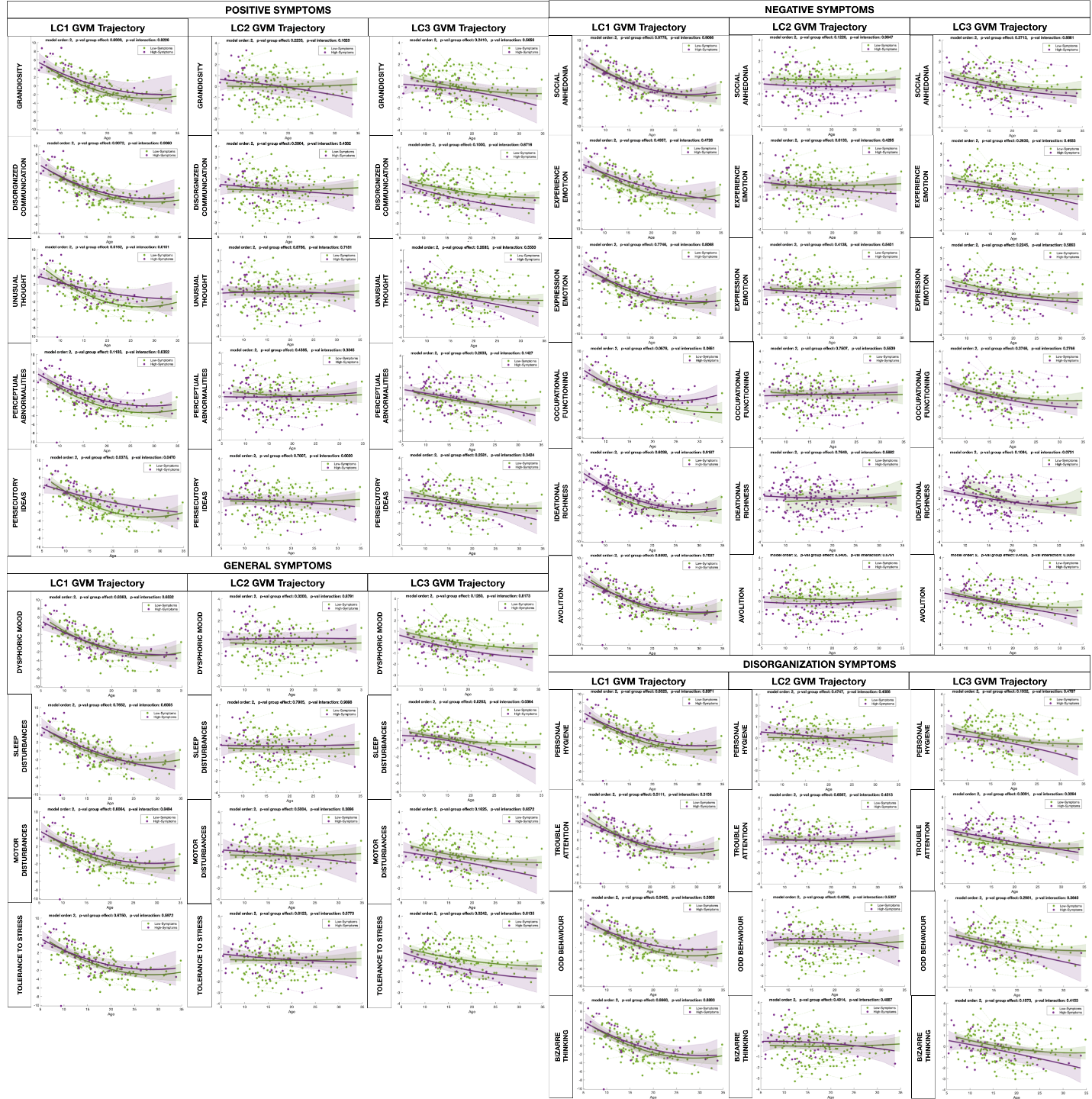

Supplementary Figure 14: Contribution of individual SIPS variables to Cortical-GMV trajectories captured by Clinical-Cortical PLSC analysis

For each significant Latent Component (LC) we derived a single multivariate Cortical-GVM score, through matrix multiplication of the PLSC loading of cortical variable with GMV values measured at each longitudinal assessment. We compared the trajectory of such Cortical-GVM scores as a function of age across individuals divided according to the intensity (>3 on SIPS Scale) of individual SIPS symptoms. Individual symptoms that were significantly associated with atypical Cortical-GVM trajectory detected by the 3 significant PLCS latent components are highlighted in purple.

1.	Mancini V, Sandini C, Padula MC, Zoller D, Schneider M, Schaer M, et al. Positive psychotic symptoms are associated with divergent developmental trajectories of hippocampal volume during late adolescence in patients with 22q11DS. Molecular psychiatry. 2019.

2.	Sandini C, Zoller D, Schneider M, Tarun A, Armondo M, Nelson B, et al. Characterization and prediction of clinical pathways of vulnerability to psychosis through graph signal processing. Elife. 2021;10.

3.	Kaufman J, Birmaher B, Brent D, Rao U, Flynn C, Moreci P, et al. Schedule for Affective Disorders and Schizophrenia for School-Age Children-Present and Lifetime Version (K-SADS-PL): initial reliability and validity data. Journal of the American Academy of Child and Adolescent Psychiatry. 1997;36(7):980-8.

4.	Reich W. Diagnostic interview for children and adolescents (DICA). Journal of the American Academy of Child and Adolescent Psychiatry. 2000;39(1):59-66.

5.	First MB GM, Spitzer R, Williams J. . Structured Clinical Interview for the DSM-IV Axis I Disorders (SCID-I). Washington, DC: American Psychiatric Association. 1996.

6.	Miller TJ, McGlashan TH, Rosen JL, Somjee L, Markovich PJ, Stein K, et al. Prospective diagnosis of the initial prodrome for schizophrenia based on the Structured Interview for Prodromal Syndromes: preliminary evidence of interrater reliability and predictive validity. The American journal of psychiatry. 2002;159(5):863-5.

7.	Overall JE, Gorham DR. The Brief Psychiatric Rating Scale. Psychological Reports. 1962;10(3):799-812.

8.	L.A ATMR. Manual for the ASEBA Adult Forms & Profiles. Burlington, VT.

: University of Vermont, Research Center for Children, Youth, & Families,; 2003.

9.	TM A. Manual for the Child Behavior Checklist/4-18 and 1991 profile: Burlington: University of Vermont Department of Psychiatry; 1991.

10.	D W. The Wechsler intelligence scale for children—third edition: administration and scoring manual. San Antonio: Psychological corporation; 1991.

11.	D. W. Wechsler adult intelligence scale-III: administration and scoring manual. San Antonio: Psychological Corporation; 1997.
